# Developing a co-produced, systems-informed, sexually transmitted infection contact tracing intervention for gay and bisexual men who have sex with men and their ‘one-off’ sexual partners

**DOI:** 10.1101/2021.07.07.21260064

**Authors:** Paul Flowers, Sarah Lasoye, Jean McQueen, Melvina Woode Owusu, Merle Symonds, Claudia Estcourt

## Abstract

**Objective:** Gay and bisexual men who have sex with men (GBMSM) bear a disproportionate burden of sexually transmitted infections (STIs). Most STIS are asymptomatic and people infected won’t know to seek care unless they are told about their exposure. Contact tracing, is the process of identifying and contacting sex partners of people with STIs for testing and treatment. Contact tracing is sometimes particularly challenging amongst GBMSM because of the kinds of sexual relationships which GBMSM enjoy. These include one-off partners who are particularly important for transmission dynamics as they contribute disproportionately to onwards transmission. The effectiveness of contact tracing interventions within sexual health are patterned by sexual-partner type. Contact tracing and management for ‘one-off’ partners is an on-going public health challenge. Low motivation amongst index patients, high resource burden on health care professionals and problems with contactability are key barriers to contact tracing. Using insights from complex adaptive systems thinking and behavioural science, we sought to develop an intervention which addressed both the upstream and down-stream determinants of contact tracing and change the system in which many inter-dependent contact tracing behaviours are embedded.

**Setting:** UK community-recruited GBMSM, stakeholders, sexual hcp, dating app providers

**Method:** Using the MRC complex intervention framework and insights from the INDEX study, a three-phase intervention development process was adopted to specify intervention content. Phase one consisted of an inter-professional and community-member stakeholder event (n=45) where small mixed groups engaged in exploratory systems-mapping and the identification of ‘hot spots’ for future intervention. Phase two used a series of focus groups with GBMSM (n=28) and interviews with representatives from key dating app providers (DAPs) (n=3) to further develop intervention ideas using the theoretical domains framework, the behaviour change wheel and the behaviour change technique taxonomy. In Phase 3 we again worked with key stakeholders expert health care professionals (HCPs) (n=5) and key workers from community-based organisations (CBOs) (n=6) to hone the intervention ideas and develop programme theory using the APEASE criteria.

**Results:** The co-produced intervention levers change simultaneously across the system within which contact tracing is embedded. Multiple change-agents (i.e., GBMSM, CBOs, HCPs) work together, sharing an overall vision to improve sexual health through contact tracing. Each make relatively modest changes that over time, synergistically combine to produce a range of multiple positively-reinforcing feedback loops to engender sustainable change around contact tracing. Agreed intervention elements included: a co-ordinated, co-produced mass and social media intervention to tip cultural norms and beliefs of GBMSM towards enabling more contact tracing and to challenge enduring STI- and sex-related stigmas; complementary CBO-co-ordinated, peer-led work to also focus on reducing STI stigma and enabling more contact tracing between one-off partners; priming GBMSM at the point of STI diagnosis to prepare for contact tracing interactions and reduce HCP and sexual health-service burden; changes to SHS environments and HCP-led interactions to systematically endorse contact tracing; changing national audits and monitoring systems to directly address one-off partner targets; delivering bespoke training to HCPs and CBO staff on one-off partners and the social and cultural context of GBMSM; DAPs active involvement in mass and social media promoting appropriate contact tracing messaging.

**Conclusion:** Our combination of multiple data sources, theoretical perspectives and diverse stakeholders have enabled us to develop an expansive, complex intervention that is firmly based in the priorities of those it will affect, and which has a solid theoretical foundation. Future work will assess if and how it will be possible to evaluate it. The resulting intervention is profoundly different than other ways of enhancing contact tracing, as it simultaneously addresses multiple, multi-levelled, upstream and social determinants of contact tracing.

## Introduction

Contact tracing and the management of those potentially exposed to infectious disease has long been a cornerstone of public health. Its primary logic is simple: once having identified someone with an infectious disease *focus* further efforts to reduce onwards transmission where transmission is most likely to have occurred. Depending on the nature of the infectious disease, its transmission routes, and our ability to treat it, further focussed efforts may include testing, treatment, a range of behaviours to reduce onwards transmission (e.g., testing, self-isolation and physical distancing) and of course more contact tracing.

Across a range of infectious diseases, the effectiveness of contact tracing efforts remain largely unknown (Fitzgerald and Bell, 1998). Because of the ubiquitous nature of contact tracing as a routine intervention in public health, evaluations of its effectiveness are relatively rare (see however Saurabh and Prateek, 2017; Swanson *et al*., 2018; Juneau *et al*., 2020) although many modelling studies do exist (Müller and Kretzschmar, 2021). However, the COVID-19 pandemic has fuelled a recent explosion of interest in contact tracing as its urgent and rapid implementation quickly highlighted major problems in the COVID-19 context. Within England, for example, contact tracing has been particularly problematic (Department of Health and Social Care, 2020; Limb, 2020). We would argue one key problematic aspects of contact tracing *per se*, as has been seen with contact tracing COVID-19 in particular, relates to the lack of attention to its psychosocial and sociocultural aspects. On a superficial level, contact tracing can of-course be thought of simply as the focusing of public health efforts on the contacts of an index patient to reduce onwards transmission. However, this somewhat deracinated perspective overlooks the complexity of the social worlds in which both the index patient and their contacts are embedded. Stepping away from the specific interactions between an index patient and their contacts opens new opportunities for intervention. A focus on the patterning of factors, which drive interactions within communities and networks that are associated with onwards transmission represents a complementary and viable alternative. In contact tracing interventions across varied infectious diseases, little attention has been paid to health inequities and the need to target and tailor contact tracing interventions to particular communities and their life circumstances (Mathevet *et al*., 2021). Equally, until recently, very little research has explored the factors shaping particular communities’ engagement with contact tracing. However, a recent rapid review highlights major barriers and facilitators to engaging in contact tracing across a range of infectious diseases. It suggests key barriers relate to: mistrust and apprehension, fear of stigmatisation, specific challenges relating to mode of its delivery (i.e., digital vs otherwise) and privacy concerns (Megnin-Viggars *et al*., 2020). In contrast, the rapid review notes that key facilitators of effective contact tracing interventions relate to the co-production of contact tracing systems, perceptions of collective responsibility, perceived personal benefit, and perceptions of the contact tracing intervention as efficient, rigorous and reliable.

It follows that if we want to enhance contact tracing interventions for infectious diseases per se and for STIs in particular, we need to systematically engage much more with their meaning and implementation, and directly address their intersection with the lives and social worlds of those they directly affect. This paper pursues this goal and seeks to begin the process pf developing an intervention to reduce onwards transmission of STIs by improving contact tracing.

### Sexually transmitted infections, GBMSM and contact tracing

Globally, more than one million sexually transmitted infections (STIs) are acquired each day, and result in serious negative health impacts (Rowley *et al*., 2019). Reducing the prevalence and rates of transmission of STIs among GBMSM has become a key area of focus for public health. GBMSM bear a particular burden of sexual ill-health. It is highly likely that the on-going, ever-transforming, social worlds of GBMSM have been shaped by the cumulative effects of homophobia, heterosexism and a lack of a salutogenic intergenerational culture. Unlike other communities in which the rhythm and cycle of childbirth, parenting, grandparenting and extended families and local communities all enable intergenerational culture to be shared across time, for GBMSM it is primarily, although not exclusively, through sex that culture is sustained. Again, although it can never be empirically demonstrated, these factors may be associated with the on-going presence of hyper sexualised sub-cultures amongst GBMSM. These subcultures are characterised by high numbers of disassortative sexual mixing which in turn are associated with higher risks of STI transmission. Arguably, increases in the use of digitally mediated sexual mixing has further contributed to these issues. Although some of the most profound behaviour change ever to have been recorded was seen in these communities (Stall et al., 2005 in response to the impact of HIV/AIDS), in recent years the incidence of STIs other than HIV has been increasing. Within the UK, GBMSM account for a disproportionate number of syphilis and chlamydia diagnoses *(Public Health England 2019;* Health Protection Scotland 2019). Addressing the rise of STI transmission among GBMSM is of increasing importance, especially given growing evidence of emerging antimicrobial resistance in key pathogens Neisseria gonorrhoea and Mycoplasma genitalium (Wi *et al*., 2017). To address this complex situation WHO have set a target to reduce the incidence of gonorrhoea by 90% globally by 2021 (World Health Organisation, 2016). They recommend combining *education, behaviour change, development of vaccines, new medications and partner management to notify and treat the sex partners of individuals diagnosed with STIs.* Partner management is a term used to describe contact tracing within the sexual health field. It is a key component of STI control by breaking chains of transmission. Contact tracing for GBMSM remains poorly understood, and effective approaches specifically for one-off sex partners in particularly are largely unknown (Wang *et al*., 2016).

### Approach to intervention development for contact tracing amongst GBMSM and their one-off partners for STIs

As part of the LUSTRUM programme of research which aims to improve contact tracing interventions for STIs & HIV we have used psychosocial and social insights to optimise and then trial a contact tracing intervention known as accelerated partner therapy (APT, see for example, (Estcourt *et al*., 2020; Pothoulaki *et al*., 2020). In typically contact tracing in the UK a health care professional asks the person with the STI (known as the ‘index’ patient) to 1) think back to all the people they have had sex with in the last 3 months, 2) inform those sex partners that they need to get tested and treated (at the sexual health clinic). Alternatively ,the health care professional offers to inform the sex partners for the index patient, usually without disclosing the index patient’s identity. In relation to LUSTRUM, and APT where the index patient takes the testing and treatment to their partners, we theorised that this particular contact tracing intervention would work well when index patients and their partners are in types of relationship where sex is likely to occur again, and affective ties motivate sexual partners to engage in contact tracing (Estcourt et al., 2021). APT, for example, even harnesses people’s on-going contact to actually deliver testing and treatment directly. In contrast, and perhaps more akin to the affective elements of much contact tracing within the recent COVID-19 pandemic, when sex is unlikely to occur again, and when there is no affect to motivate contact tracing and enable further focussed public health efforts, contact tracing initiatives are much harder to gain traction. Actively being part of contact tracing is only associated with costs (e.g., self-isolation in COVID). For GBMSM and their one-off partners the costs of engaging in contact tracing with one-off partners stem the lives and social worlds of those affected, for example, cultural factors such as the on-going negative impacts of homophobia and heterosexism which in turn shape STI-related stigma and negative attitudes towards sex (Flowers et al., 2021). At the individual-level of particular GBMSM, issues such as reputational damage and perceptions of culpability and blame are also important considerations (Flowers, Duncan and Frankis, 2000: Flowers et al., 2021). Equally, from the perspectives of those who provide health services, it can be hard to justify the considerable resources required to achieve good contact tracing outcomes in harder to reach contacts, despite them contributing disproportionately to community transmission.

Given the importance of one-off sexual contacts to onward transmission and the difficulties in achieving good PN outcomes in this group we did an intervention development study that aims to improve contact tracing amongst GBMSM and their ‘one-off’ partners. Particularly mindful of the psychosocial and sociocultural factors that shape contact tracing *per se* and our insights into the specifics of contact tracing with ‘one-off partners’, we were keen to use the best available approaches to intervention development. Recent insights into the science of intervention development from the INDEX study (O’Cathain *et al*., 2019) and the updated MRC Complex Intervention guidance (Medical Research Council, 2019) highlight the centrality of working with stakeholders from the start of the intervention development process. They also highlight the need to theorise interventions and their causal mechanisms in relation to the specific contexts in which they will be delivered. Recent studies in the science of intervention development also demand more focussed attention on long term sustainability and attempts to achieve intervention implementability early on within the intervention development process. In this way, we can avoid the oft reported ‘implementation gap’ associated with apparently successful interventions, showing promise within trials, later failing to translate into routine practice (Murray *et al*., 2010; May, 2013).

In relation to using theory within intervention development we used a a combination of theoretical approaches. Complex adaptive systems (CAS) perspectives are heralded as being particularly useful approaches to seemingly intractable public health problems (Wilson, Holt and Greenhalgh, 2001; Rutter *et al*., 2017). They are particularly good at enabling an overview of a problem and generating *novel* solutions to potential interventions that are not limited by our typical constrained and linear understanding of cause and effect. They encourage us to think wholistically and systemically and deal with complexity head on. These approaches focus our attention on ways to intervene which are not overly simplified and instead highlight the need to address upstream (i.e., distal) and often multi-facetted causal influences (e.g. interdependent and multi-levelled drivers which shape system behaviour in ways that are challenging to reduce). As such we sought to use these systems perspectives at the start of our intervention development process. However, we would argue that although CAS perspectives are useful for imagining new ways of intervening, they are far less useful for actually operationalising the intervention ideas they can generate. Therefore, we also drew upon a range of theories and approaches from implementation science that assist with the clear specification and operationalisation of intervention ideas. (for more details see the intervention development protocol (McQueen *et al*., 2020). In this way we drew upon a range of other theoretical frameworks that focus on implementation per se and ways of changing implementation behaviour.

### Aim

To develop a co-produced, systems-informed, multi-level, contact tracing intervention for GBMSM with bacterial STIs and their “one-off” partners.

### Research Questions

1. What did diverse stakeholders tell us about where best to intervene within the system that drives contact tracing?
2. What can we learn through behaviourally informed qualitative work to suggest particular evidence-based and theoretically-informed future intervention content
3. What is the final suggested intervention content after further stakeholder engagement?

## Methods

A full account of the design and details of each phase of the research is detailed within the initial protocol (McQueen *et al*., 2020). Here we provide a brief overview.

### Design

Overall a three-phase design was used (See Figure 1). The first phase consisted of a major stakeholder event, an appraisal of the literature, and discussions with experts in the field. Initial ideas for intervention components generated at this phase were explored and developed further within Phase 2 and Phase 3.

> **Phase 1** *What did diverse stakeholders tell us about where best to intervene within the system that drives contact tracing?*

**Figure 1:**
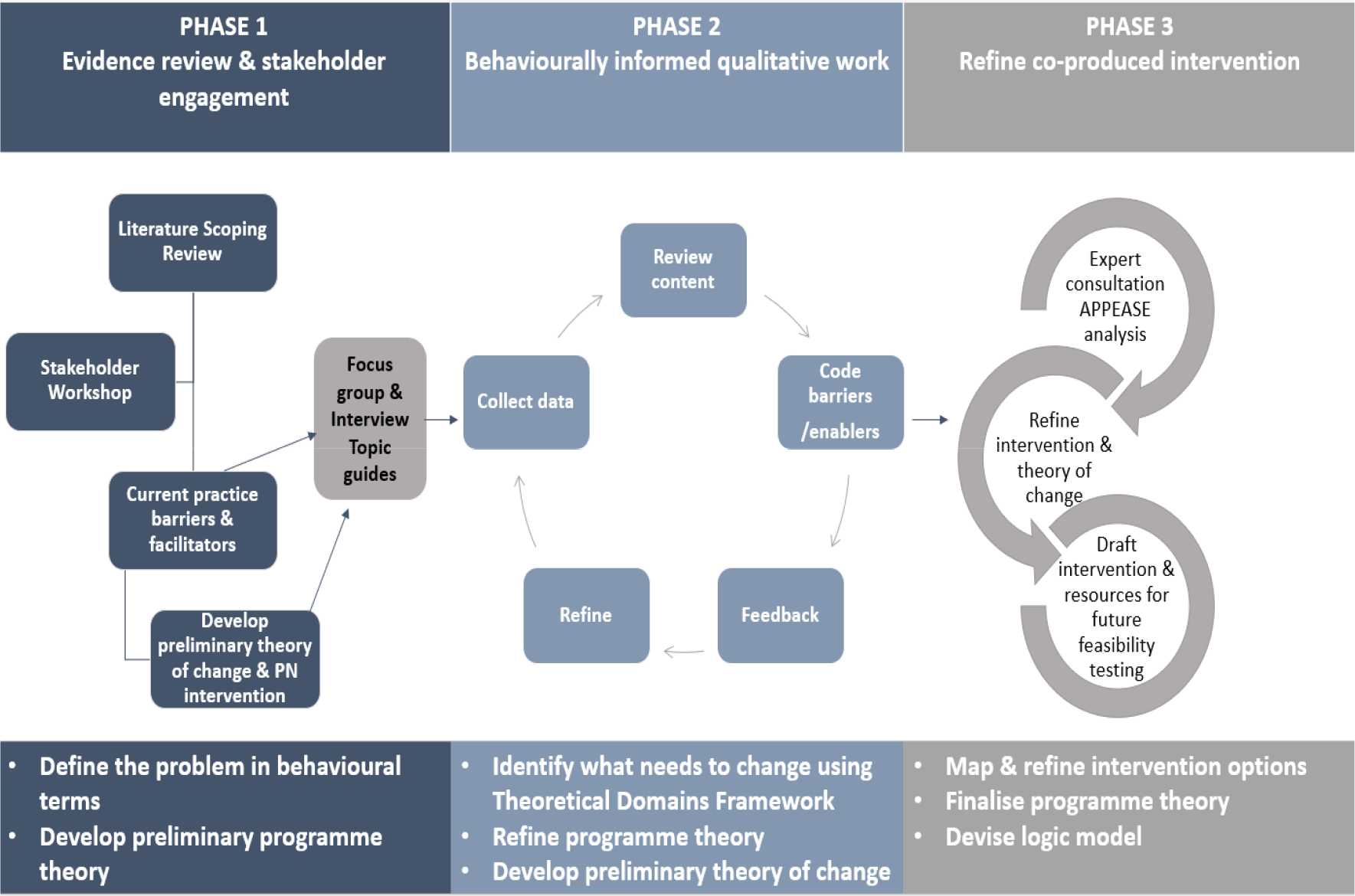
Diagram presenting the components and process of each phase of the intervention development study.

The stakeholder event aimed to work with important stakeholders to map the system which shapes contact tracing and begin to identify potential areas and methods of future intervention.

### Data collection

Within the stakeholder workshop a series of small mixed groups were convened and tightly facilitated to ensure the problem of contact tracing for one-off partners was seen wholistically and explored from multiple vantage points simultaneously. Small groups consisted of GBMSM, those working in CBOs, sexual health care professionals, service commissioners, sexual health researchers and those working in the dating app industry. Topics of discussion included the overall system which drives contact tracing, and an exploration of the full range of key organisations, communities, professionals, individuals, social processes, dynamics, and interactions involved and connected within the system. Throughout, in the context of GBMSM and their one-off partners, participants were encouraged to consider what currently worked to facilitate contact tracing and conversely what did not work well and why. Participants were also asked to think expansively and creatively to consider if improvements to contact tracing for GBMSM and their one-off partners were possible, and if so where and with whom this might take place and why they thought this might be important. Equally participants were asked to note where and why improvements were unlikely to ever be possible. Discussions were not audio recorded. Data was collected in the forms of visualisations from participants during systems-mapping and detailed notes were collected by scribes during facilitated group-discussion

### Data Analysis

The stakeholder workshop data analysis focussed on systematically appraising the visualisations, detailed notes and sticky labels. PF and JMcQ considered *how* participants mapped the system shaping current and potential contact tracing and extracted a series of heterogeneous potential ‘*hot spots’* where future intervention could usefully focus. An informal rapid review of the available published literature was also conducted at this time to assess the nature and relative success of existing interventions in this field. These ideas were examined in relation to workshop insights. PF and JMcQ began the process of considering potentially useful intervention ideas for future interventions to improve PN with MSM and their one-off partners. Through presentations and discussions with the wider team they were able to formulate working hypotheses about potentially useful intervention ideas. In this way a very basic and provisional programme theory was being formulated.

> **Phase 2** *What was learned through behaviourally informed qualitative work to suggest particular evidence-based and theoretically-informed future intervention content?*

We further explored the nascent intervention ideas with samples of people who would be affected by the intervention. Analytic tools from implementation science were used to understand barriers and facilitators to the implementation of the emerging intervention ideas developed from Phase 1 and to specify ways of optimising these ideas to secure an acceptable, feasible, sustainable future intervention.

### Data collection

Focus groups with GBMSM, health care professionals (HCPs) and Dating App providers (DAPs) were planned to further develop initial intervention ideas. This sampling strategy and the topic guides for both the interview and focus group participants were informed by the initial intervention ideas generated by stakeholders. Participants were asked to discuss the emerging intervention ideas, whether and how they could be implemented, and the barriers and facilitators to implementation. All interview and focus group data were digitally recorded and transcribed.

### Data Analysis

The data were analysed firstly to asses broad acceptability and whether participants thought the implementation of the intervention idea was actually feasible. Subsequently deductive thematical analysis was then undertaken to detail barriers and facilitators to implementing the intervention ideas deemed acceptable and amenable to change. Further data analysis then consisted of using the Theoretical Domains Framework (TDF) to theorise important barriers to the uptake or delivery of intervention ideas. Then the Behaviour Change Wheel (BCW) approach, incorporating the Behaviour Change Technique Taxonomy (BCTT), was used to specify potential intervention content in granular detail. Analyses were led by JMcQ and audited by GV and PF - both trained within these approaches (Michie, van Stralen and West, 2011; Michie *et al*., 2013; Michie, Atkins and West, 2014; Atkins *et al*., 2017).

> **Phase 3** *What is the final suggested intervention content after further stakeholder engagement?*

Although our original aim had been to work with a similarly diverse range of interprofessional stakeholders that had contributed to Phase 1, this was no longer possible because of the impact of COVID 19 which precluded the conducting of research which involved HCPs as participants.

### Data collection

Instead, the project relied on working with staff from community-based organisations (CBOs) and working with sexual health experts from within the research team itself (primarily HCPs). This phase involved a series of four virtual workshops where participants were presented with both an overall high-level account of the emerging intervention ideas, as well as specific and granular detail generated during Phase 2. Phase 3 participants were asked to consider specific intervention ideas in relation to the APEASE criteria (APEASE=Affordability, Practicability, Effectiveness and cost-effectiveness, Acceptability, Side-effects and safety, Equity) and comment on whether content should be kept, modified or abandoned. All data was digitally recorded and transcribed.

### Data Analysis

JMcQ and PF appraised data collected within these virtual workshops and iteratively worked with the data to identify content from the developing intervention which could be kept, abandoned or modified. Colour coding (green, amber, red) was then applied to the detailed intervention content outlined in Phase 2 to result in a final list of intervention content (Appendix 1).

## Results

### Phase 1: What did diverse stakeholders tell us about where best to intervene within the system that drives contact tracing?

In November 2019 the stakeholder event recruited 45 diverse participants representing a broad spectrum of agents within the system that drives contact tracing. These included GBMSM with no professional interest in the topic (20%), public health / sexual health service commissioners (17%), representatives of community-based organisations (9%), Health advisors and nurses (17%), sexual health researchers (11%), medical consultants from sexual health (20%) and representatives from DAPS (6%). A substantial minority of the stakeholders were both professionals and GBMSM.

Together, stakeholders highlighted how drivers of effective contact tracing were complex and distributed across a complex system. Figure 2 provides a schematic overview of the systemic drivers effecting contact tracing, collating stakeholder input from the initial engagement event. The drivers of contact tracing are multi-levelled and considerably inter-dependent. There is a clear balance of connected drivers that both enable and constrain contact tracing.

**Figure 2.**
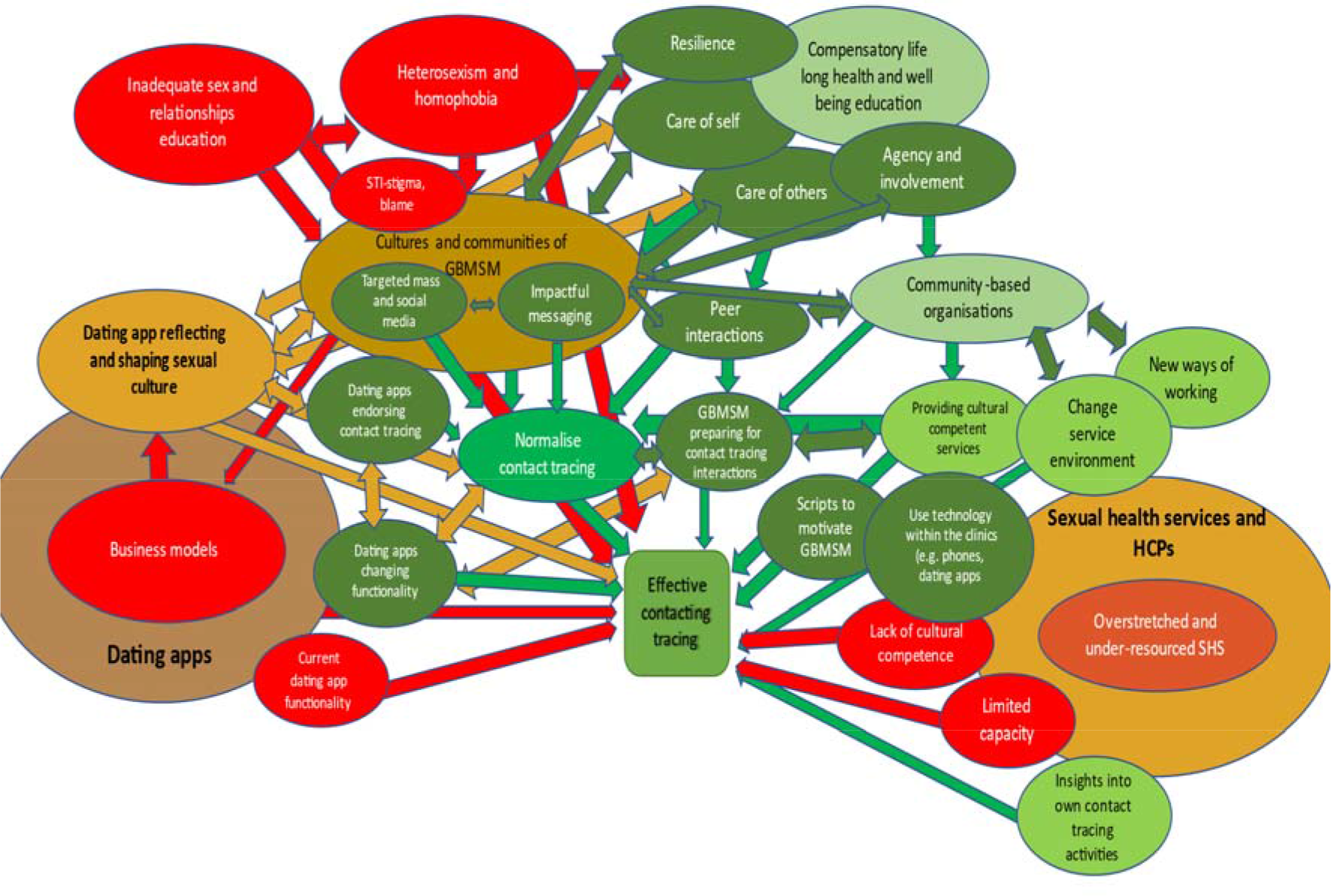
Schematic overview of the diverse systemic drivers effecting contact tracing, including pathogenic (red) and salutogenic (green) influences collated from the initial stakeholder event.

Stakeholders described how many critically important drivers of effective contact tracing tended to be distal to the actual act of notifying partners (e.,g., the legacy of inadequate sex and relationship education). Current contact tracing approaches with their primary focus on proximal determinants (e.g, using index patient affect to motivate patient referral) were understood to be necessarily limited when applied to one-off partners (e.g., practical and resources-related barriers) particularly within the age of digital dating apps (e.g., blocking)

Table 1 below summarises the findings of the stakeholder workshop and what we learned about the drivers of contact tracing.. It should be noted that, when compared to Figure 2, the tabularisation of these issues minimises the sense of how interlinked and interconnected the drivers of contact tracing are. A full narrative account of the drivers is available within supplementary file A.

**Table 1.** The multi-levelled and systematic drivers of contact tracing.

|  |
| --- |
| **The distal drivers of contact tracing** |
| *Negative drivers (factors limiting contact tracing with one-off partners)* |
| The socioeconomic context of sexual health service provision |
| Overstretched sexual health services |
| The business models of dating apps |
| Elements of contemporary and historical culture |
| The on-going impact of heterosexism |
| The on-going impact of homophobia |
| History of pathologizing and blaming GBMSM |
| Legacy of inadequate sex and relationship education |
| High levels of sex and STI-related stigma |
| *Positive drivers (factors facilitating contact tracing with one-off partners)* |
| A distributed approach to improving drivers of contact tracing that did not over burden any single stakeholder |
| The agency and involvement of GBMSM throughout |
| Collective and co-ordinated efforts across the wider system |
| Saturate the system with clear signals about cultural value of contact tracing |
| Working with communities of GBMSM to enhance existing salutogenic aspects |
| Use existing assets: presence and history of existing norms about contact tracing, community resilience in relation to HIV |
| Harness peer interactions to drive improved contact tracing |
| Use all available infrastructures (websites, dating apps, broad range of health services) |
| Compensatory sexual health education |
| New cultural norms about contact tracing |
| New cultural expectations about contact sharing |
| Co-produce mass and social media intervention elements to change GBMSM cultures and norms |
| Ensure dating apps promote and endorse norms to facilitate contact tracing |
| Change dating app functionality and features |
| **The proximal drivers of contact tracing** |
| *Negative drivers (factors limiting contact tracing with one-off partners)* |
| Sexual health services experienced as unwelcoming to GBMSM |
| Interactions between HCPs and GBMSM experienced as judgemental and unhelpful |
| *Positive drivers (factors facilitating contact tracing with one-off partners)* |
| Opportunities for HCPs to reflect on current and optimal practice |
| Data driven approaches to understand current success and failure |
| Opportunities to share good practice within and across services |
| Training to ensure HCP cultural competence in relation to GBMSM |
| Scripts for HCPs to motivate GBMSM to engage in contact tracing |
| Work with GBMSM to anticipate and prepare for contact tracing interactions within sexual health services |
| Enhanced use of digital technology within contact tracing interactions (smart phones, dating apps) |
| Work with key people organising sex parties, sex clubs for GBMSM |
| Work around contact tracing within public sex environments |

#### The selection of intervention ideas

Using the APEASE criteria (i.e., Affordability, Practicability, Effectiveness/cost-effectiveness, Acceptability, Side-effects/safety, Equity) and consideration of which elements of the system were actually amenable to change, Phase 1 ended with an appraisal and selection of key areas to explore in greater depth and detail during Phase 2 (see Figure 3 for an overview). Our stakeholders’ system mapping had highlighted that future interventions must be *multi-levelled* (i.e. addressing both distal and proximal determinants of contact tracing). It also suggested that they must be *multi-agent* in order to share the burden of activity across the whole system whilst simultaneously normalising contact tracing in new ways. These core principles have the potential of realising sustainable change by re-organising the system itself to in turn improve contact tracing. These key principles were taken forwards to the subsequent phases.

**Figure 3.**
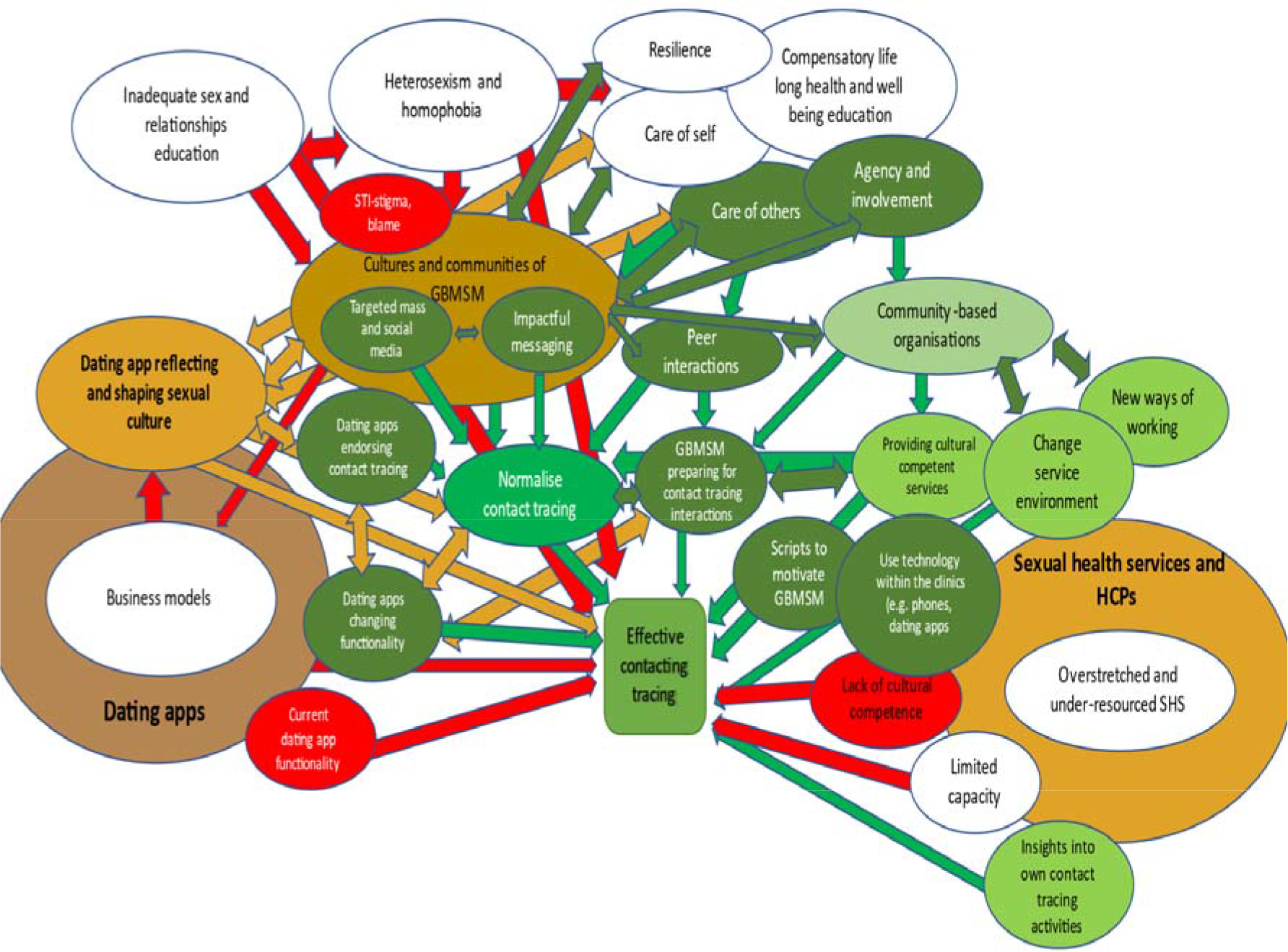
Selection of key areas to examine further within Phase 2

Although acknowledging their fundamental importance in shaping contact tracing we chose not to further examine the following because they were not amenable to change within the intervention being developed here: inadequate sex and relationships education, the need for compensatory lifelong health and well-being education for GBMSM to foster better care of the self and the other, focussed efforts to reduce heterosexism and homophobia, attempts to build the resilience of GBMSM, the business models of the dating app industry, the economic drivers of health services challenges that stretch sexual health services. Although we had chosen to remove these elements as they were not particularly amenable to change, we were also mindful of the potential of these elements to re-emerge within Phase 2 and Phase 3 as we learned more from our participants and stakeholders.

We chose to focus on how communities of GBMSM, dating apps providers and sexual health services could work together in new ways and which complementary and co-ordinated activities could mutually reinforce each other. Such activities, included, targeted mass and social media with impactful messaging that changed aspects of gay culture that constrained contact tracing yet celebrated aspects that enabled it, dating app endorsement of contact tracing and hosting of media messaging, changing app functionality to enable GBMSM to take part in contact tracing, changing peer interactions to exact social influence echoing the key messages of targeted mass and social media, enhancing the ways HCPs worked with GBSMS though using technology within contact tracing interactions, through providing consistent and multiple messages through interactions and changes to the service environment to make contact tracing more salient, through providing culturally competent services able to support GBMSM and their contact tracing with one-off partners, through using motivational and culturally competent scripts to enable contact tracing with one-off partners and through using data driven approaches to enable services to learn more about their relative success and failures in contact tracing through (e.g. audit, monitoring, service reviews).

### Phase 2 What was learned through behaviourally informed qualitative work to suggest particular evidence-based and theoretically-informed future intervention content?

The selected preliminary intervention components from Phase 1 were re-considered and further developed through a series of focus group discussions and interviews with key stakeholders. Critically, because of the impact of COVID-19 on HCP and NHS staff it was not possible to collect data from diverse HCPs as had been intended. As such the proposed intervention ideas generated in Phase 1 did not benefit from the critical appraisal of HCPs. For some of the intervention ideas relating to the SHS this was a particular loss. Developing new ways of working for sexual health clinics, for example, or addressing issues of cultural competency, harnessing insights into current good practice, were unable to be further developed by diverse HCPs in this phase. Focus groups with GBMSM were facilitated in Glasgow, London and Leeds. Two further focus groups were facilitated virtually (n=28). Telephone interviews were held with DAPs (n=3).

Overall within this phase, and bearing in mind the caveats concerning the lack of input from sexual health services and HCPs, there was on-going support for key intervention elements from Phase 1.

In the paragraphs below we illustrate how participants thought about acceptability and whether intervention ideas were amenable to change. The participants also indicate key barriers and facilitators to their implementation.

Participants agreed with the principle of needing to work across the system in different ways at the same time ‘all these different tools, all these different ways, you know, to make it easier to just inform people’ (GBMSM virtual 1) as was the need not to overburden any particular stakeholder ‘we need to find a way that we’re not heavily just relying everything on health care professionals, but also creating a culture of ownership, responsibility, and awareness, in the LGBT community. (GBMSM virtual 1). Some of the later data collection events also stressed the timeliness of the project and talked about how COVID-19 had done a huge amount of groundwork for the current project ‘what makes this conversation, particularly easy for everyone to understand now, is the experience of being locked down, the test and trace discussion around Covid-19 and being able to illustrate how that connects to STIs. So, everyone is in a different place right now than if we’d been having this conversation 12 months ago. So, it’s linking to that Covid-19 experience, must make selling this concept a lot easier’.(DAP2). Against this positive support for the intervention ideas as a whole, there was some mention of extant challenges. It was felt important to acknowledge the on-going and longstanding dynamics that negatively shape cross-stakeholder partnership work ‘And I think that, you know, as we talk about contact tracing and then we think about the sort of history of partner notification, there’s a really bad relationship there based on behaviour of public health in terms of gay men and gay men sexuality [….] There’s a history there and they’re not approaching this from a sort of neutral standpoint….bias and homophobia. I think that there are ways that we can also sort of work more collaboratively on some of the messaging and work more collaboratively on, you know, frankly repairing some of that relationship damage (DAP1)

In relation to what GBMSM said about changing their cultures and communities to normalise contact tracing there was widespread agreement and enthusiasm for progressing these areas of future intervention. The pre-COVID focus groups in particular highlighted a lack of knowledge and understanding about contact tracing amongst some GBMSM. They also highlighted a lack of consistent norms about what men should and could do about notification. For some GBMSM this was the norm but for others it clearly wasn’t. However there was a clear sense of the need to address these issues within future intervention. ‘I think what we need, first of all, is more visible campaigning, promotion, advertising, social media, that it becomes cool to say it, it just becomes cool, it makes a person who brings this up more cool, than somebody who doesn’t bring it up. So what we need is some kind of social media campaigning that normalises people discussing openly with each other’. (GBMSM Virtual1). There was also consensus and agreement that peer-focussed work between GBMSM was important ‘the facts are that people have sex, everyone should notify each other if they get something. That’s It.’ (GBMSM Glasgow).There was clear agreement to progress ideas such as targeted mass and social media including impactful messaging to normalise contact tracing.

Across the focus groups there was considerable heterogeneity of experience about contact tracing. From the pre-COVID focus groups in particular, there was a clear acknowledgment that it was not always a central feature of GBMSMs culture and that there was a lack of consistent knowledge about what contact tracing is, and what should happen around it, ‘If everyone had heard the same information, we’d probably be notifying our partners a lot more, but because of inconsistency in the information being given, it just isn’t there. Not everyone’s at the same level of access to it’.(GBMSM Leeds). Filling the basic knowledge gap around contact tracing amongst GBMSM was thought to be an important first step in progressing contact tracing interventions ‘I think normalising the words [about contact tracing] will normalise talking about it, will normalise telling each other about it. (GBMSM, Glasgow). Across the focus groups here was a shared sense of the high value and cultural acceptance of earlier STI and HIV mass media interventions. Discussion of these effective interventions focused on ideas about potential essential requirements for future contact tracing interventions, for example: ‘it’s got to be a very clever strapline, and a very kind of, immediately known, immediately remembered, and everyone knows exactly what it’s about. So yeah, you’re right, it’s got to be something short, but clear’. One focus group went as far as outlining a potential key message ‘Three Ts – test, treat, tell. (GBMSM, Virtual focus group 2). Equally, when considering the knowledge gaps around contact tracing, participants talked about how important it was to model what contact tracing was like: ‘maybe showing a video of what it’s like to tell someone, actually seeing something like that, I think that would be…actually, if we’re talking about the fear of it, let’s actually have something that shows that experience’. (GBMSM, Virtual 1).

In relation to what DAPs told us about implementing the earlier intervention ideas, there was consensus across the interviews regarding the need for working in partnership across the system, there was also strong agreement with the need for culturally appropriate social and mass media. New suggestions emerged concerning associated merchandising ‘do something that people are willing to wear’. There was also a clear acknowledgement of the need for targeted messaging that genuinely celebrated the diversity of GBMSM communities and used culturally appropriate figures ‘porn stars and sex workers and their perspective on sexual health issues as opposed to a professional talking about it in a dry sense of the word. (DAP2). DAPs highlighted the need to think about the diversity of GBMSM within interventions and issues of visual representation ‘I feel like every single gay person no matter where they fit in the community has to be able to look at that and realise that, okay, this is affecting me as well. So, it can’t just be, you know, white guy who basically dresses in H and M head to toe and is saying, oh, you should tell your partner [inaudible 16.09], you know, head to toe in tattoos and blah blah’.

DAPs also highlighted the constraints they felt as small organisations with limited capacity to work beyond their primary remit as businesses ‘We are a team of four probably I think it’s everyone, I mean we’re a small organisation so there’s only so many hours in the day’ (DAP2). However, there was support for the idea of signing up to a code of conduct relating to contact tracing but also an acknowledgement of its practical problems ‘I mean the challenge would be agreeing what are the standards that we’re going to offer to users and what’s the technology required to get there’. In contrast to the relative enthusiasm for being involved in messaging and media interventions, changing functionality to enable contact tracing was not seen as a viable option, not only would the process be complex but ‘Its around the right to block someone. And I think that would kind of violate it. I’d probably say most dating apps would say no’.

In relation to what GBMSM said about SHS and innovations in contact tracing (‘new ways of working’) many highlighted the potential of innovations within services, where future changes could ‘start to feel you’re being treated more like a human being’ (GBMSM, Virtual focus group 2). For some GBMSM there was appreciation of the pressure facing many services and how this limited innovations in contact tracing. Time invested in one patient, receiving intense motivational counselling for contact tracing for one-off partners, for example, was understood to essentially take time away from others ‘that means somebody else is waiting 20 minutes more just to go in and get tested’ (GBMSM, London). However, they also perceived other opportunities within the services that could be harnessed for contact tracing ‘you’re waiting in the waiting room, the TV screen, they can do it (educate) that way as well, while you’re waiting you just look at the screen for helpful messages tips’ (GBMSM, London). Equally, some of the interaction time within services could be reduced if GBMSM prepared and shared information before arriving in the service, saving what was perceived to be valuable HCP time, ‘if they sent you a link or something where you can send them back the number or a Grindr profile or whatever’. (GBMSM, London). Overall however, GBMSM felt that HCPs within some services could do more to engage GBMSM to do more in relation to contact tracing ‘I think, the attitude of the people at those more progressive, and welcoming clinics, are a bit more supportive in terms of the actual, like, practical need for support, to do the disclosure bit’ (GBMSM, Virtual focus group 2)

Our analysis went beyond the analysis of acceptability and barriers and facilitators to implementation outlined above. Because of the size of our comprehensive analyses of data collected in this phase we do not present the details of our theorisation of them using the TDF. Equally, we cannot show here how we used the BCW and the BCTT to generate highly specific evidence-based and theoretically informed ways in which the Stakeholders initial ideas could be operationalised in ways that systematically reduced barriers to implementation yet harnessed perceived facilitators. However, for the interested reader the full analyses underpinning these aspects of Phase 2 are presented within the supplementary files (Tables 1-7).

As a very overarching summary of this work, our deductive analysis generated 17 barriers and 15 facilitators. We generated 83 potential intervention functions (Environmental restructuring, education, enablement, persuasion, modelling, training and incentivisation) and 141 potential BCTs. The full analyses underpinning this Phase of the research are available as supplementary files (Tables 1-7).

In relation to broad changes where our participants noted intervention ideas were not acceptable and/or not amenable to change the overall span of intervention ideas was reduced. This primarily included the abandoning of attempts to change dating app functionality and the use of technologies within sexual health services. Figure 4 shows an overview of the remaining intervention ideas that were taken forwards to Phase 3.

**Figure 4.**
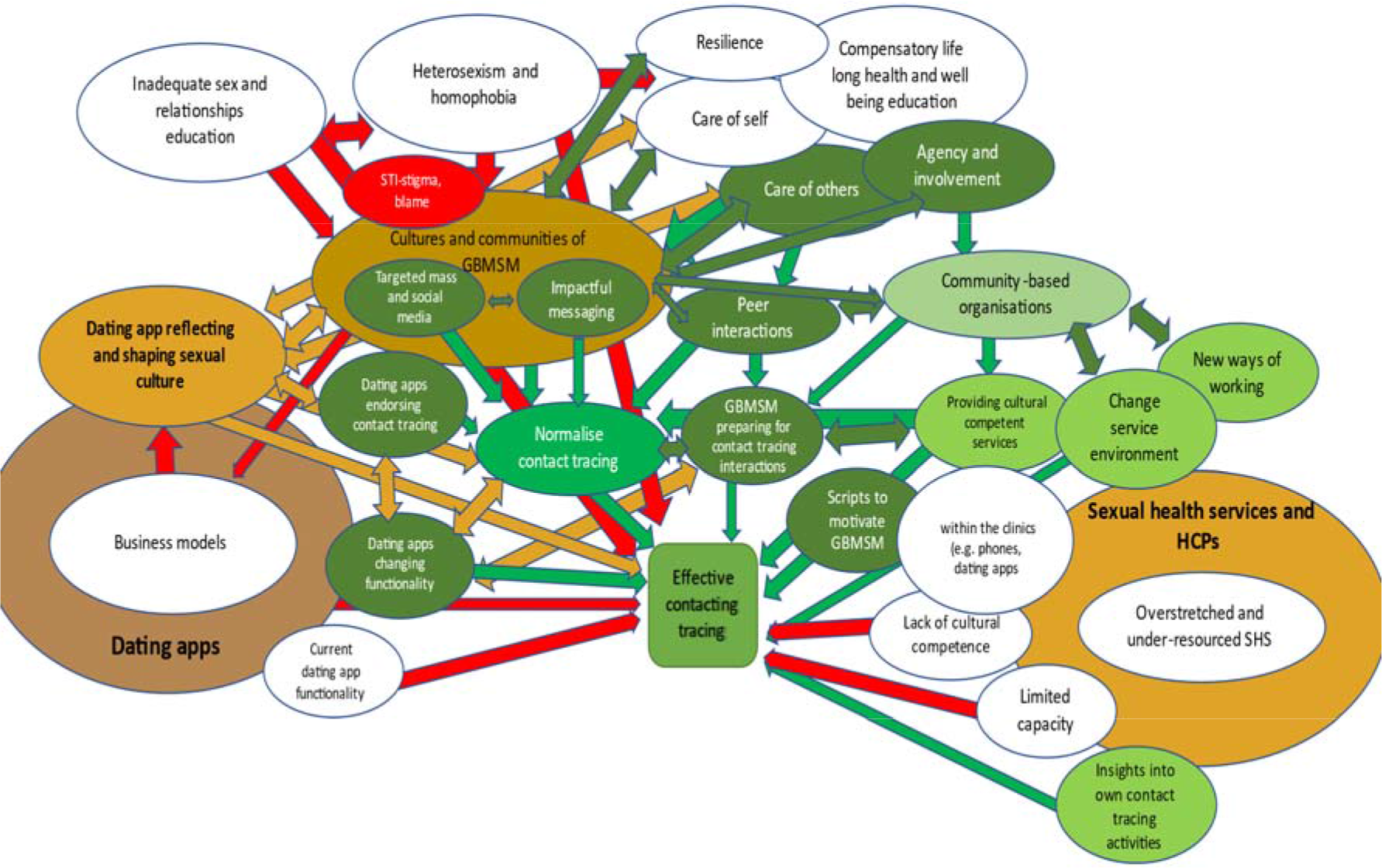
An overview of intervention ideas after detailed consideration of how to implement them and taken to Phase 3

### Phase 3 What is the final suggested intervention content after further stakeholder engagement?

A second large stakeholder engagement event mirroring that at the start of the project had originally been planned to take place, wherein intervention components developed and specified at the end of Phase 2 would be finalised for inclusion. However, due to the outbreak of COVID-19, this approach was no longer possible due to reduced stakeholder availability and the severe limitations of online facilitation for complex multi-stakeholder engagement. Consequently, Phase 3 was re-assessed, and an alternative scaled-down approach to refining the intervention’s final components through stakeholder engagement was agreed. This consisted of two stakeholder groups with community-based organisations with expertise in the field of sexual health and GBMSM (n=9, representing four organisations) and two stakeholder events with diverse sexual health clinicians drawn from the wider LUSTRUM research team (n=5). Using the materials generated at the end of Phase 2 these stakeholders firstly discussed the general principles shaping the emerging intervention. Subsequently, they discussed the range of individual intervention components that Stage 2 had generated in detail. Throughout the APEASE criteria were used to facilitate rounded discussions about whether intervention ideas should be kept, modified or rejected.

In relation to the idea of working in targeted mass and social media interventions with impactful messaging, all stakeholders explored earlier intervention ideas such as ‘*test, trace, tell and treat*’ with other messaging such as ‘*Identify, Inform, Invest, Improve*’ (Expert clinicians, 2) wherein people Identify their contacts, Inform their contacts, Invest in a check-up and together this Improves your sexual health and those of the community. Echoing Phase 2 community-based stakeholders here also emphasised the need to represent the diversity of GBMSM communities and ensure materials depicted a diversity of experiences and perspectives. There was agreement about the need to educate all GBMSM about contact tracing ‘*there should be like some sort of educational awareness in terms of telling people or educating them how to respond if somebody tells you about, if they’ve been diagnosed with an STI’ (CBO stakeholder group1)* and indeed to challenge STI-related stigma and blame ‘*extinguishing blame I think is definitely the central thing that we need to out’ (CBO stakeholder group2)*. Community-based stakeholders noted the need for positive focus ‘*the positive effects of like feeling a sense of responsibility to your community’ (CBO stakeholder group 2)*. There was also support for the idea of comparing two alternative story lines, one depicting effective contact tracing and one showing an example of contemporary blame culture, ‘*I think any campaign should talk about why there’s a benefit to you on both sides. So, it might be the quote is, ‘that dirty bastard!’ and you can turn round and say, actually, ‘I think it was great, because he got in contact with me, and he was looking after my health, so I was able to get treatment’. So, you switch it, you know, it’s kind of a solutions focus, you make the negative into a positive. And you make it about how it’s benefitting you and that person, and vice versa’ (CBO stakeholder group, 2)*.

From the community-based organisations there was consensus regarding the need to empower GBMSM to be at the heart of driving new interventions to improve contact tracing. There was support for the idea of peer driven work in this regard although caveats around appropriate funding to support such activity were clearly expressed. It was felt that volunteers were not an appropriate resource for these activities. Clear ideas about working with and celebrating community action were expressed ‘*one thing that really frustrates me more than anything about when we make advances, like PREP and U=U, is you’ll have the clinicians shouting how we’ve done it, you’ll even have NGOs shouting about we’ve done it, and to be frank, the only people that have actually done this and brought down the HIV rate and things like that, is the community, because they have taken action to do that. And for me, that’s the really important thing to get across, that any intervention is, bring the community in, because they’re the actual people that make that intervention work’ (CBO stakeholder group,1)*. In turn galvanising community action was seen as a potential mechanism that could overcome some of the enduring barriers to delivering contact tracing for one-off partners amongst HCPs and sexual health services. Expert clinicians felt that GBMSM actively asking for help from HCPs in relation to seeking assistance with protecting their communities was seen as a very powerful way of activating overstretched HCPs (e.g., paraphrasing some of the recent community activism around PrEP, ‘*I want PN now’*).

Expert clinicians also agreed about the value of working with SHS and HCPs to learn from insights into their own successes and failures. Indeed it was felt that this mechanism not only could deliver on-going improvements but that it could also assure a consistency of contact tracing approaches across varied and deeply heterogeneous services. However, it was agreed that nationally co-ordinated guidance was a probably a more effective and efficient means to achieve these ends than attempts to host ‘insight-sharing’ initiatives within cross professional cross service platforms for example. Finally, all stakeholders agreed on the need to improve HCPs cultural competencies and provide services which could accommodate and facilitate contact tracing for GBMSM and their one-off partners. However expert clinicians acknowledged this might require approaches such as using secret shoppers and giving feedback in non-confrontational ways. Other suggested approaches included making more use of community-based organisations ‘*the third sector, have a great opportunity to build a culture where [contact tracing] is acceptable, and I’m not necessarily sure that the statutory sector can do that*’ (CBO)

Key intervention elements that expert clinician stakeholders suggested should be abandoned included modifying the clinical environment to reinforce contact tracing messages. Stakeholders with expertise in SHS suggested this may not work for the diverse patients using the service from a range of backgrounds and for very diverse reasons

Final granular recommendations concerning *how* the acceptable interventions ideas that have been through multiple APEASE processes could be implemented are presented within Supplementary files (Tables 1-7). In addition, we provide a narrative summary of our agreed core principles and components and this can be found in Appendix 1.

## Discussion

The current study sought to follow contemporary guidance about intervention development (O’Cathain et al., 2019) and move the field forwards by systematically working with a range of stakeholders to develop a novel contact tracing intervention for GBMSM with STIs and their one-off partners that could perhaps overcome the multiple and multi-level barriers that have prevented the development of effective interventions in this area of contact tracing to date. As such this paper makes a novel contribution because of its use of diverse stakeholders, its use of diverse theoretical lenses and its focus on the psychosocial and socio-cultural determinants of contact tracing in addition to those that are more service oriented. This approach contrasts sharply with the recent glut of poorly conceived, atheoretical, app-based interventions that attempt to address contact tracing in the context of COVID-19 primarily through the development of Apps (Colizza et al., 2021). Overall, within the study we have worked with diverse stakeholders to see the problems and potential solutions to contact tracing with one-off partners. Across three phases of the project, we have iteratively honed a long-list of potential intervention ideas into a much shorter list of intervention ideas that are both acceptable and implementable and if they were to be operationalised may stand a chance of delivering sustainable change.

Our study has foregrounded the insights of key stakeholders and understands the drivers effecting contact-tracing amongst GBMSM and their one-off partners to relate to a complex and interconnected system. Through iterative data analysis, participant knowledge gleaned from system-mapping, exploration, and critical interviews and discussion, we have been able to refine the core components and content of a wholistic future intervention. To summarise refined intervention components focus on collective action across the system and final recommendations underscore the collaborative cross-sector nature of future interventions.

Our analysis suggested critical areas for future intervention included enabling and empowering GBMSM communities to do more about contact tracing and STIs with their one-off partners. Agreed activities in this area included peer-led initiatives, mass and social media with targeted impactful messaging. The content of such messaging could challenge STI-related stigma and raising awareness of contact tracing amongst GBMSM whilst also articulating norms and scripts of how to tell one-off partners about their risk of exposure and how to react appropriately. Working in partnership with CBOs and DAPs throughout this endeavour should enable the seamless promotion of these intervention elements across the social and sexual environments in which GBMSM meet. In addition to moderating some of these wider upstream and cultural determinants of contact tracing, it was also felt that enabling GBMSM to prepare themselves for contact tracing interactions with HCPs was also a good idea. This capitalised on the agency and responsibility of GBMSM. It was felt that this enabled them to seek support and help from SHS and HCPs rather than expecting overstretched HCPs and SHS to deliver this kind of work. DAP in-put, HCP guidance.

In relation to remaining uncertainties pertaining to the intervention and potential next steps it is important to acknowledge that the current study has only begun the process of intervention development and has successfully outlined potential intervention content. Our major investment in these first steps of effective intervention development were designed as a solution to the on-going problem of rapidly conceived interventions failing to gain traction with the communities and services they are intended to work within (e.g, Chalmers et al., 2014; Walugembe et al., 2019). As a result of this emphasis, key questions relating to how the emerging interventions could be funded, how it could be evaluated and the feasibility of successfully collecting and analysing such data all remain to be answered.

### Limitations and strengths

The outbreak of COVID-19 in the UK in early 2020 severely affected the timeline and planned operations of the key phases of this study. Disruptions to Phase 2 in particular impacted negatively on the project as whole. This negativity relates primarily to the loss of HCP perspectives in appraising and further developing intervention ideas within Phase 2. Given the centrality of diverse perspectives and the idea of problem solving from multiple vantage points simultaneously, the loss of the projects ability to systematically hone intervention ideas using sexual health services and HCPs perspectives remains a limitation. Our work with expert clinical stakeholders from within the project team did of course help compensate for this loss. As a result, some aspects of potential intervention ideas concerned with SHS and HCPs have only been illuminated by GBMSM adopting a ‘patient perspective’ and our expert clinical stakeholders within Phase 3. As such issues such as ‘using more technology such as mobile phones within clinical interactions’, ‘using techniques such as motivational interviewing or scripts to assist with contact tracing with one-off partners’, ‘the need for more culturally competent services’, and ‘ways of overcoming limited capacity’ could potentially have been assessed and modified differently if we had been able to recruit diverse HCPs working within the field. Similarly, although we did manage to recruit some DAPS to the project, our attempts at recruitment struggled in the context of this sector adapting to the impacts of COVID-19. Again, this represents a limitation to the overall design. A final limitation relates to Phase 3 and the impact of COVID on our stakeholder work. Ideally, this would have mirrored Phase 1 with an interactive mixed small groups of professionals working with the intervention ideas. Once more, given COVID-19 restrictions this was not possible. Further limitations include our sole reliance on qualitative approaches, if time and resource had permitted a more expansive mixed methods design would have enhanced the project, tools like using the Delphi approach could have been used.

Strengths of the project include its systematic use of stakeholder input which has ensured the intervention ideas are grounded in the priorities of those the potential intervention will affect. Our use of diverse stakeholders ensured that the potential intervention did not either overburden one part of the system and indeed that it did not shunt the problem elsewhere within the system. Other key strengths included the studies pluralistic approach to using theory. Where complex adaptive systems theory was useful, we used it to see the nature of problem the intervention was trying to solve differently. This enabled us to see the importance of intervening upstream as well as well as downstream. However, where complex adaptive systems theory is weak, we used behaviour change theory to systematically understand and then specify ways to ensure the implementation of the potential intervention ideas our stakeholders had generated. The overall approach has provided a number of lenses and levels in which to see the intervention we have generated. This varies from visual overviews (Figure 2-5), narrative summaries (appendix) to very granular accounts of intervention content (Tables 1-7).

**Figure 5.**
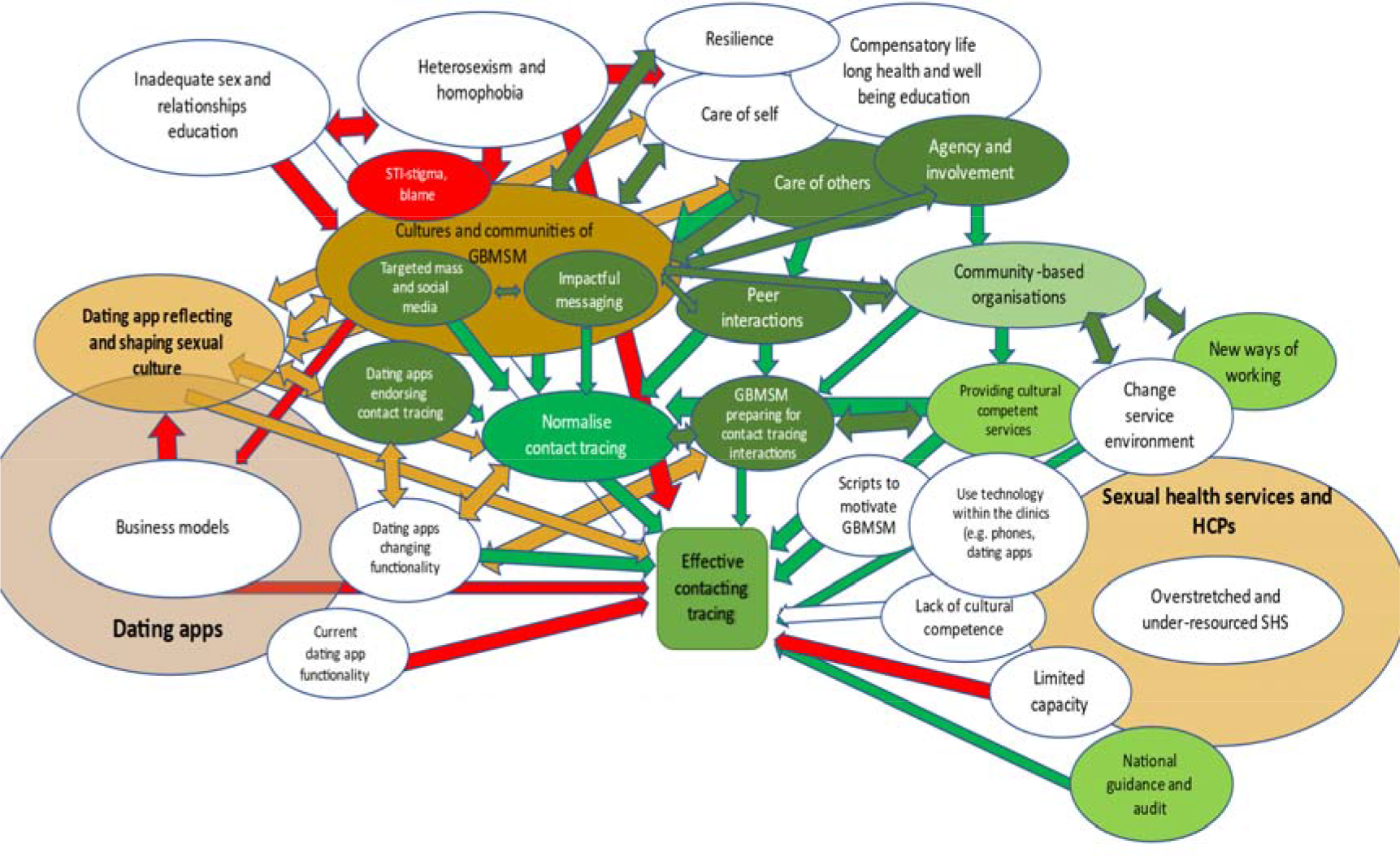
An overview of intervention ideas after final appraisal and modification with stakeholder groups

## Conclusion

We have undertaken a process of complex intervention development which foregrounds the insights of key stakeholders and understands the drivers effecting contact-tracing amongst GBMSM to be a complex and interconnected system. Through iterative data analysis, participant knowledge gleaned from system-mapping, exploration, and critical interviews and discussion, we have been able to refine the core components and content of a holistic intervention. Final recommendations underscore the collaborative cross-sector nature of potential interventions to intervene in this area. Our suggested intervention ideas are multi-level (upstream and downstream intervention elements), multi-agent (GBMSM, CBOs, HCPs, DAPs) and consist of co-ordinated and complementary activities including mass and social media elements, peer-led initiatives, the priming of individual GBMSM to prepare for interactions within SHS, and HCP training assuring a standard level of cultural competency as well as adequate national guidance and monitoring to enable SHS to appraise and constantly improve their contact tracing outcomes.

## Data Availability

Given confidential nature of the qualitative data it is not available. Data supporting analyses are at reasonable requests

## Appendix 1: Narrative summary of suggested intervention components

### Core principles

Diverse ‘change agents’ (communities, organisations, professions, individuals) should work
together to change the system that drives effective contact tracing for one-off sexual partners. This will require new infrastructure (e.g., meetings, processes) that enable a shared and fair approach to improving such contact tracing
The ‘burden’ of the intervention should be shared across the system with each change agent using
its particular expertise.
Simultaneously address both the *distal* and *proximal* determinants of effective contact tracing for
ne-off sexual partners
Expect system change to be slow and challenging but sustainable in the long-run

### Core intervention components (as summarised and narrativised from data tables)

What did we learn from GBMSM about **implementing changes relating to the specific content of targeted mass and social media interventions** to normalise contact tracing with one-off partners through impactful messaging? (Table 1)

❏ GBMSM should be targeted for interventions to educate them on contact tracing, change behaviours and promote effective contact tracing for all former-partners, especially one-off partners. Mass media interventions should be developed to promote contact tracing for GBMSM with one-off partners. Messaging about contact tracing and one-off partners should be targeted, short, punchy, easy to remember. Phrases and terms that build on recent developments from COVID and contact tracing might be helpful:-4T’s Test, Treat, Trace, Tell. This kind of work will enable MSM to tell more one-off partners. Messaging should always be branded with reputable organisations, and preferably co-produced with, and being visibly inclusive of, a diverse range of MSM. The content of these messages should educate men about the value and importance of PN, and the consequences of blame and stigmatisation in relation to STIs.
❏ Public health messages and materials should be developed to educate GBMSM on contact tracing. In order to implement these intervention ideas a national stakeholder or steering committee, including experienced HCPs, CBOs, GBMSM and other relevant representatives, should co-produce and decide on a range of consistent and clear intervention messages. Intervention messages and materials should be tailored to specific STIs, distributed and made available widely across virtual and physical locations where men meet, and focus on increasing awareness of contact tracing amongst GBMSM. Intervention messages should be clearly branded with reputable, culturally-appropriate organisations, and should build on recent learning from COVID-19 public health messaging. Intervention messages and materials should provide an example of how to tell a one-off partner about their heightened risk of STI acquisition, highlighting the health and wider consequences of not undertaking contact tracing. This could feature a short film or animation. This works as an example to follow and use in practice, and also helps to set norms concerning what to do and expect from others. Examples would be based on input from experienced HCP, other staff and peer MSM

What did we learn from GBMSM about **implementing peer-focussed work between GBMSM to endorse contact tracing with one-off partners** (Table 2)

**Table 1.**
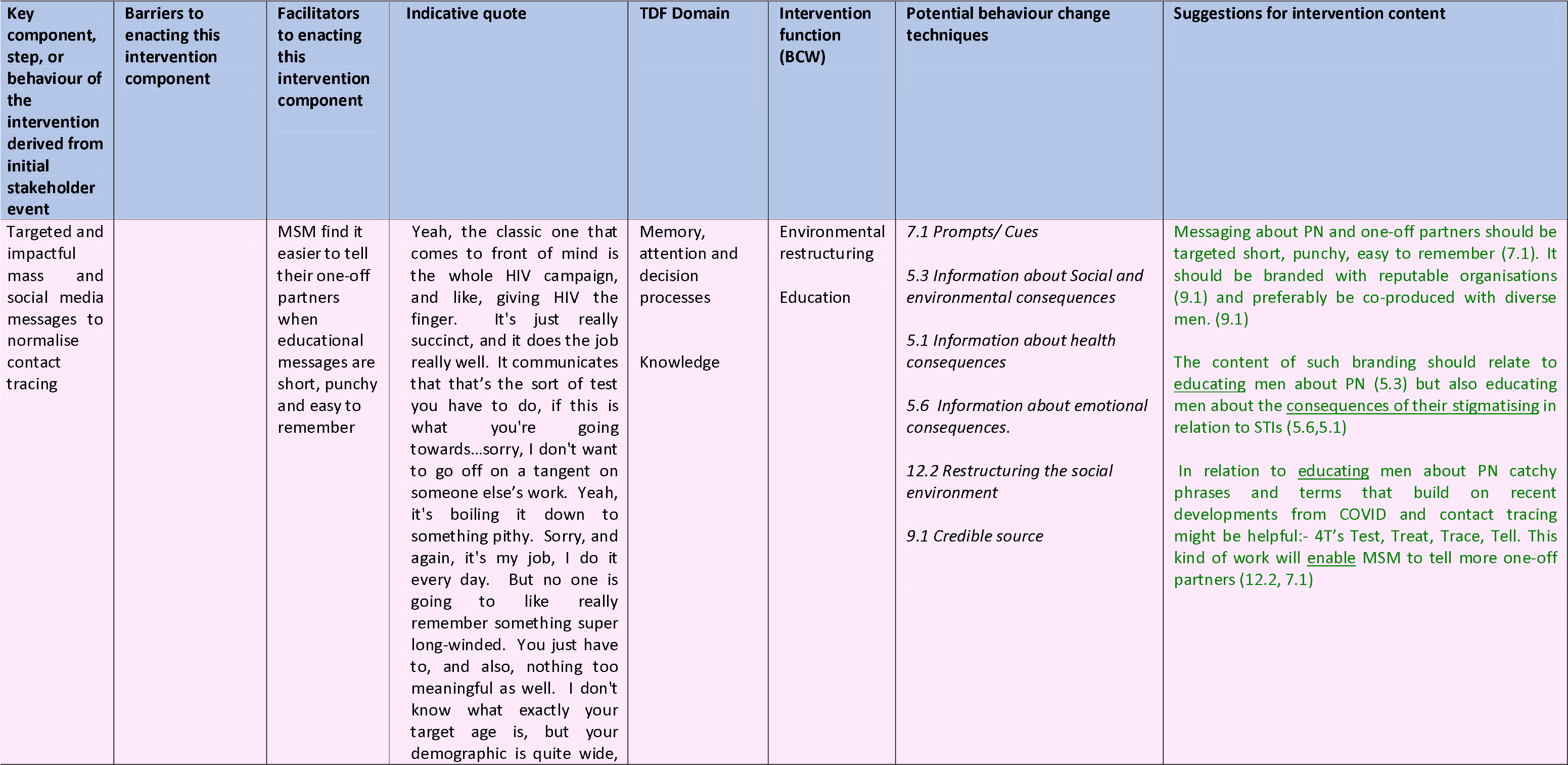
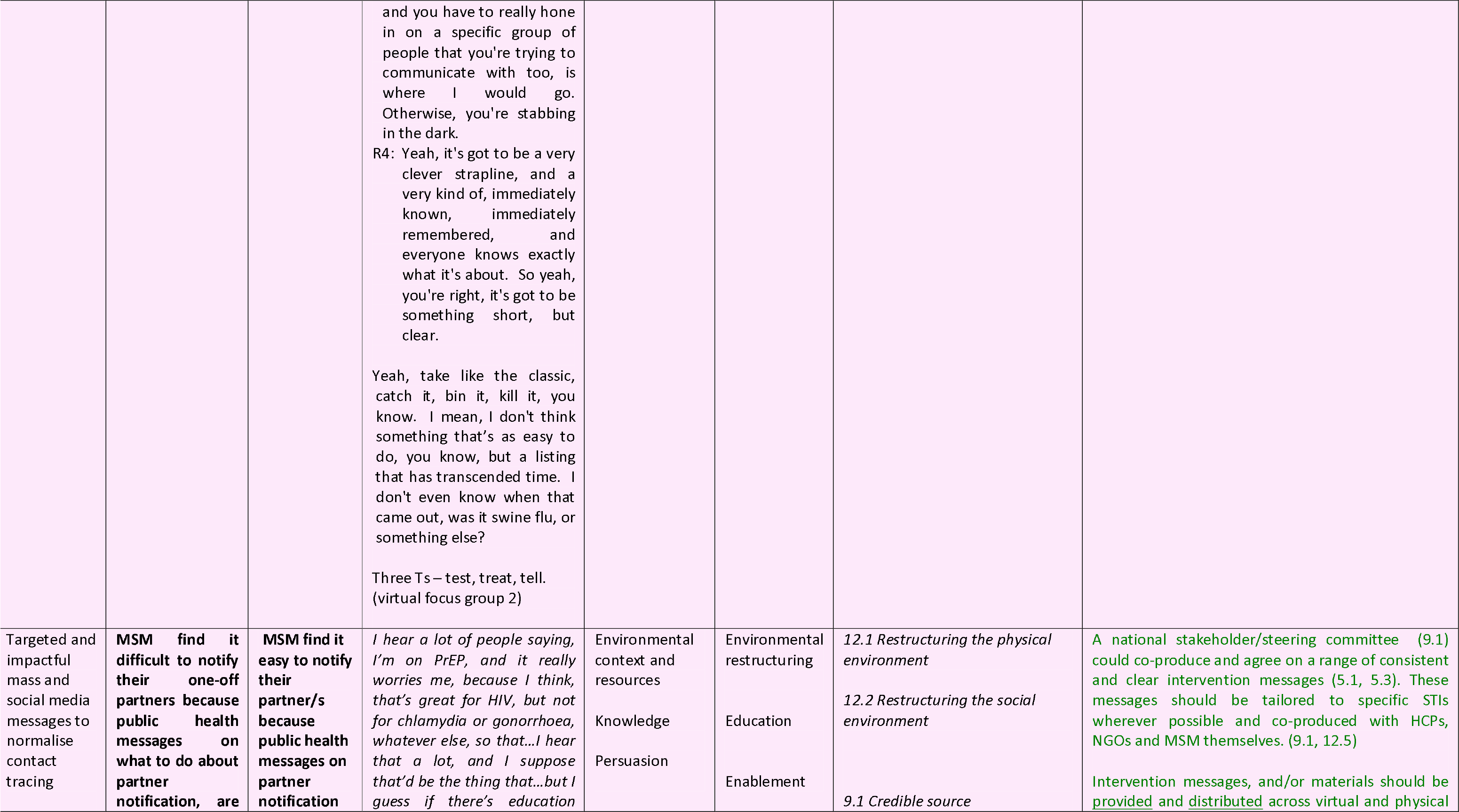
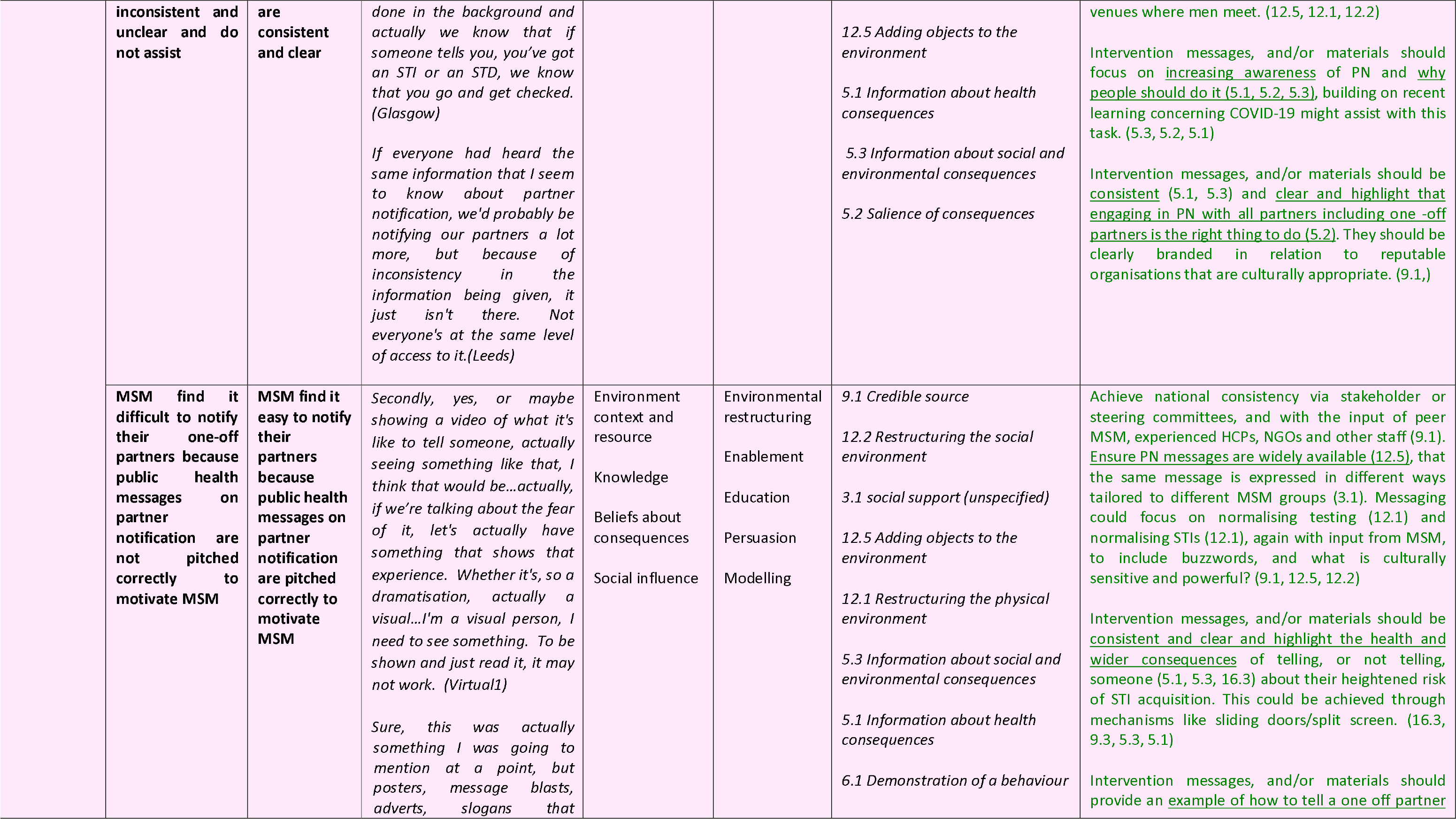
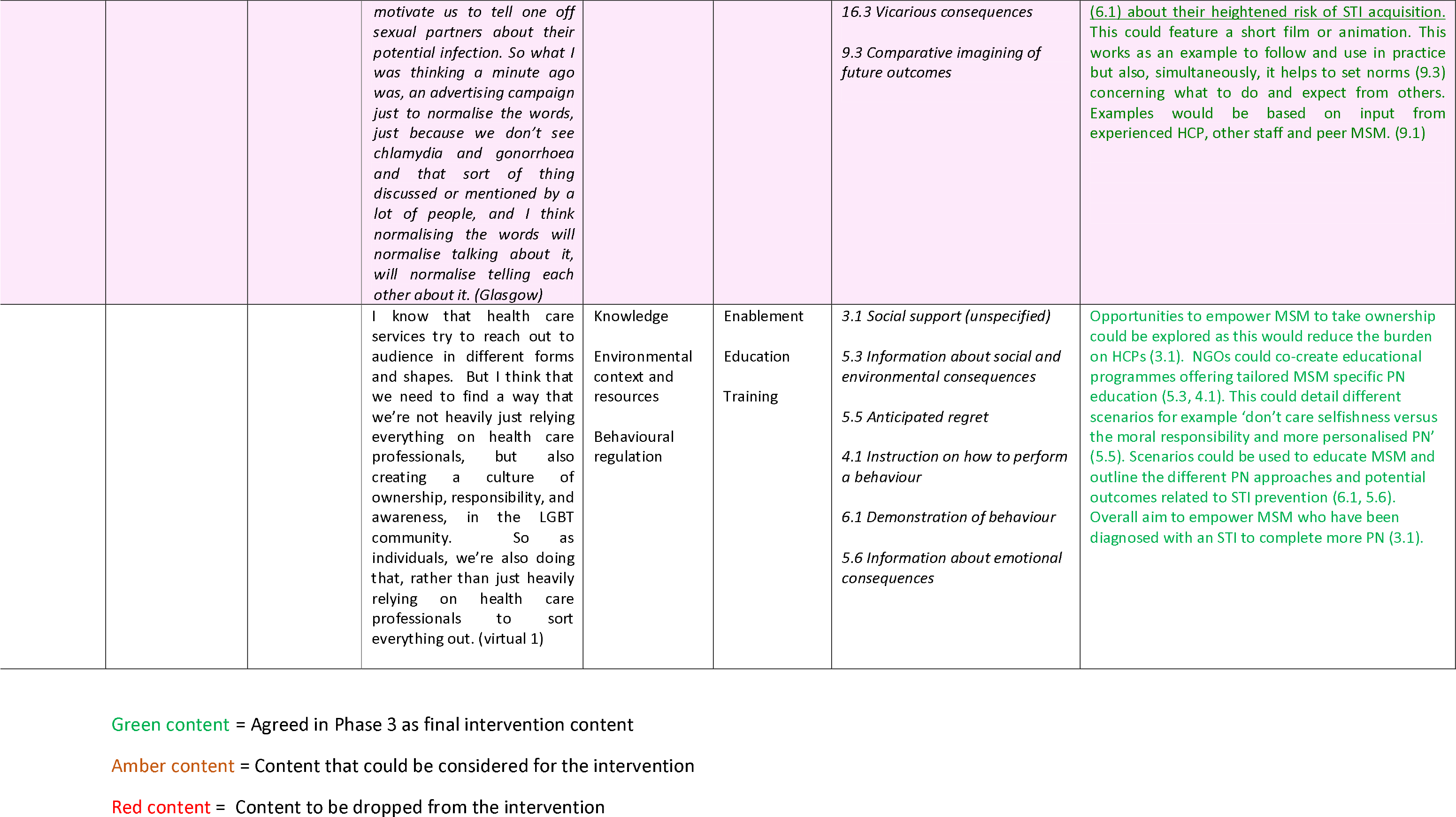
What did we learn from GBMSM about implementing changes relating to the specific content of mass and social media interventions to normalise contact tracing through impactful messaging?

**Table 2.** What did we learn from GBMSM about implementing peer-focussed work between GBMSM to endorse contact tracing with on e-off partners

| Key component, step, or behaviour of the intervention derived from initial stakeholder event | Barriers to enacting this intervention component | Facilitators to enacting this intervention component | Indicative quote | TDF Domain | Intervention function (BCW) | Potential behaviour change techniques | Suggestions for intervention content |
| --- | --- | --- | --- | --- | --- | --- | --- |
| MSM educate each other on STIs and PN | **3.1 MSM find it difficult to 'educate each other' because school-based sex education lacks key factual information about partner notification and is not tailored to MSM** | **3.2 MSM find it easy to 'educate each other' because school-based sex education covers key factual information about partner notification and is tailored to MSM** | *It should just be normal, it (the education) should just be the facts are that people have sex, everyone should notify each other if they get something. That's it. I think it should just be normal and if it is factual like that and not kind of sexual, it's about sex but not sexual, then it just becomes the norm (Glasgow)* | Environment context and resource<br>Social influence<br>Knowledge<br>Skills | Environmental restructuring<br>Education<br>Training<br>Enablement<br>Persuasion | 9.1 Credible source<br>8.3 Habit formation<br>4.1 Instructions on how to perform a behaviour<br>3.2 Social support (practical)<br>8.6 Generalisation of target behaviour (i.e. PN with all partners including one-offs)<br>5.3 Information about social and environmental consequences<br>6.1 Demonstration of a behaviour<br>7.1 Prompts/cues<br>12.2 Restructuring the social environment | To compensate for the failure of mainstream education to provide men with adequate levels of sex education (12.1, 3.2) including issues of partner notification, there is a clear need for intervention materials and messaging to <u>focus on issues of PN amongst men</u> (4.1, 5.3). These materials should be branded as having been <u>developed by reputable organisations (9.1)</u> (e.g., NGOs) and preferably be co-produced by men themselves enhancing the normative content. (9.1, 8.6)<br>It may be possible and <u>useful to focus on the power of peers (9.1)</u> to teach each other about PN; what PN is and why it should be done and why it should be done for one off partners in particular. (3.2, 5.3, 9.1, 12.2). Peers could be used to educate MSM about the moral side of PN and how it is the right thing to do (9.1, 3.2, 5.3).<br>Intervention messages, and/or materials, training should provide <u>an example, suggestions of wording (4.1), of how to tell a one off partner about their heightened risk of STI acquisition</u>. These would work as examples for others to follow and use in practice (7.1, 6.1), but also, simultaneously, helps to set norms concerning *what to do* and *what to expect*<br>*from others. (8.3))*<br>Education programmes within schools <u>should include the importance of regular STI testing (12.2) and partner notification, with case studies, scenarios and tips on how to do this well (4.1).</u> This should be part of an inclusive sex and relationship educational package informed by a variety of stakeholders, including young MSM (6.1, 9.1, 8.3) |

Green content = Agreed in Phase 3 as final intervention content
Amber content = Content that could be considered for the intervention
Red content = Content to be dropped from the intervention

❏ A range of peer-led work should be designed and delivered to enable GBMSM to capitalise on existing community norms supportive of contact tracing and also reducing norms which present barriers to contact tracing. Community-based organisations may be best placed to deliver this work. Peer-led initiatives will need funding and are unlikely to be viable through volunteering routes.
❏ Training or up-skilling opportunities should be offered to GBMSM on how to talk with other GBMSM about contact tracing and reducing STI-related stigma. Approaches could draw upon the recent activism seen around PrEP and focus this on contact tracing or partner notification (PN), for example, ‘I want PN now’. Such approaches could work with classic models of peer education (e.g., key opinion leaders), or ‘social influencers’ within the social media to change descriptive norms (what people think other people do) and injunctive norms (what people think they should do) around contact tracing with one off partners. Such peer influence could also clearly model how and why to do contact tracing for all partners.
⍰ Educational materials, including enabling approaches to peer education, should be developed for GBMSM on STIs and the value and importance of contact tracing. To compensate for the failure of mainstream education to provide men with adequate levels of sex education including issues of contact tracing, there is a clear need for a range of intervention activities (e.g. peer interactions), materials and messaging to focus on re-framing issues of contact tracing amongst men. These materials should be developed and branded by reputable organisations such as CBOs and be co-produced alongside diverse GBMSM themselves, enabling culturally appropriate content. Educational materials should focus on celebrating the power of peers to teach each other about contact tracing; what it is, why it should be done, how it should be done and why it should be done for one off partners in particular.
❏ Educational materials should provide an example, suggestions of wording for how to tell a one-off partner about their heightened risk of STI acquisition. These would work as examples for others to follow and use in practice but also, simultaneously, helps to set norms concerning what to do and what to expect from others. Educational materials should also directly address the social and sexual cultures of men and reduce perceived negative consequences of talking about STIs to enable more peer conversations about contact tracing to take place.

What did we learn from GBMSM about **using the social media to endorse contact tracing with one-off partners** (Table 3)

**Table 3.** What did we learn from GBMSM about using the social media to endorse contact tracing with one-off partners

| Key component, step, or behaviour of the intervention derived from initial stakeholder event | Barriers to enacting this intervention component | Facilitators to enacting this intervention component | Indicative quote | TDF Domain | Intervention function (BCW) | Potential behaviour change techniques | Suggestions for intervention content |
| --- | --- | --- | --- | --- | --- | --- | --- |
| Peer interactions and Social media approaches |  | 5.1 MSM find it easy to notify one-off partners as social media campaigns, and advertising promote PN making it normal practice for MSM to discuss STIs openly | *I think what we need, first of all, is more visible campaigning, promotion, advertising, social media, that it becomes cool to say it, it just becomes cool, it makes a person who brings this up more cool, than somebody who doesn't bring it up. The person taking part in discussing with the first potential partner is something cool, rather than uncool. So what we need is some kind of social media campaigning that normalises people discussing openly with each other. (Virtual1)* | Social Influence<br>Reinforcement<br>Knowledge<br>Beliefs about consequences | Education<br>Persuasion<br>Environmental restructuring<br>Modelling<br>Enablement<br>Incentivisation | 9.1 Credible source<br>6.2 Social Comparison<br>6.3 Information about others approval<br>6.1 Demonstration of a behaviour<br>12.5 Adding objects to the environment<br>3.1 social support (unspecified)<br>5.1 Information about health consequences<br>5.3 Information about social and environmental consequences.<br>12.2 Restructuring the social environment | Social media could be used within multimodal or multimedia interventions <u>to change norms (8.3, 6.3)</u> amongst men concerning PN and <u>promote particular norms</u> about engaging in PN with one off partners. This messaging would detail the diverse consequences of PN for all. (5.1, 5.3, 6.2, 12.2)<br>It may be possible <u>to recruit social influencers (9.1)</u> from the social media to promote PN and harness their networks to promote key messages. (5.1, 5.3, 12.5, 12.2)<br>Social media could be used to show people taking part in PN (6.1) with one off partners and in this way <u>provide examples of how to do it</u> and reinforce norms that doing it is the right thing to do. (5.1, 5.3, 6.3)<br>Using social networks and dating app sites <u>to exchange, repeat and 'share' messaging (12.5)</u> concerning PN with one off partners will reinforce the message and the norm of doing PN. (3.1, 5.1, 5.3) |
|  |  |  |  |  |  |  | MSM receive focused consistent messaging about PN and the health and community impact (5.1, 5.3) from key sources such as HCPs, NGOs, dating sites, (9.1) physical and virtual hook up sites. These complimentary messages should be consistent, tailored to MSM sexual cultures and reinforced by a range of stakeholders. (5.3, 5.1) |

Green content = Agreed in Phase 3 as final intervention content
Amber content = Content that could be considered for the intervention
Red content = Content to be dropped from the intervention

Social media should be utilised as a key component in targeting GBMSM. Social media could be used within multimodal or multimedia interventions to change norms amongst GBMSM and challenge stigma surrounding contact tracing, and promote particular norms encouraging engagement in contact tracing with one-off partners. Social media could be used to show people contacting one-off partners and, in this way, provide examples of how to do it and reinforce norms that doing it is the right thing to do. Social networks and dating app sites could be used to exchange, repeat and ‘share’ messaging concerning contact tracing. GBMSM should receive focused, consistent messaging about contact tracing and the health and community impact of effective contact-tracing from key sources such as HCPs, CBOs, dating apps, and physical and virtual hook up sites. Messaging would detail the diverse consequences of PN for all. It may be possible to recruit social influencers from social media to promote PN and harness their networks to promote key messages

What did dating app providers tell us about implementing changes relating to the endorsement of contact tracing within their public facing infrastructure? (Table 4)

**Table 4.**
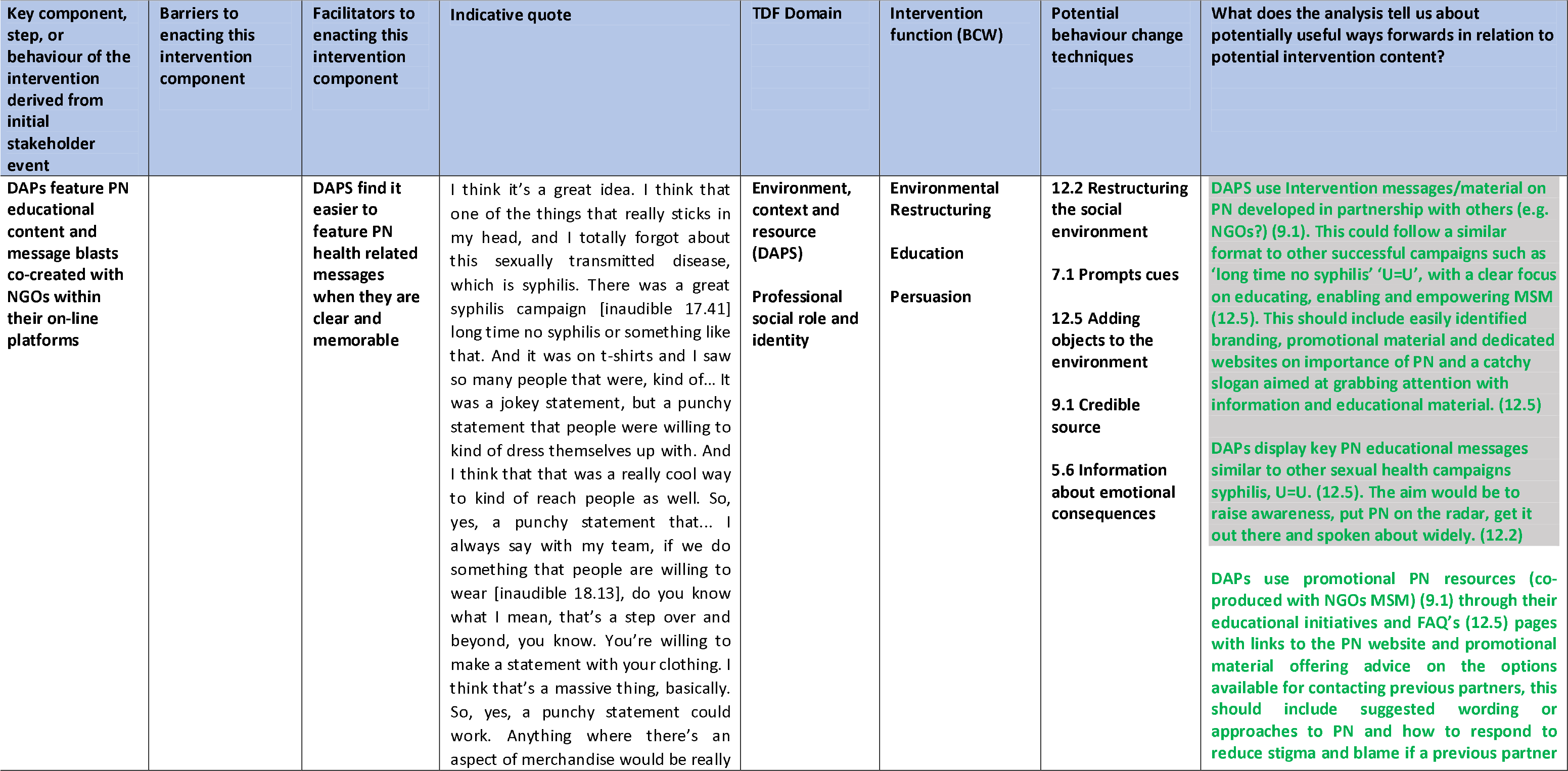
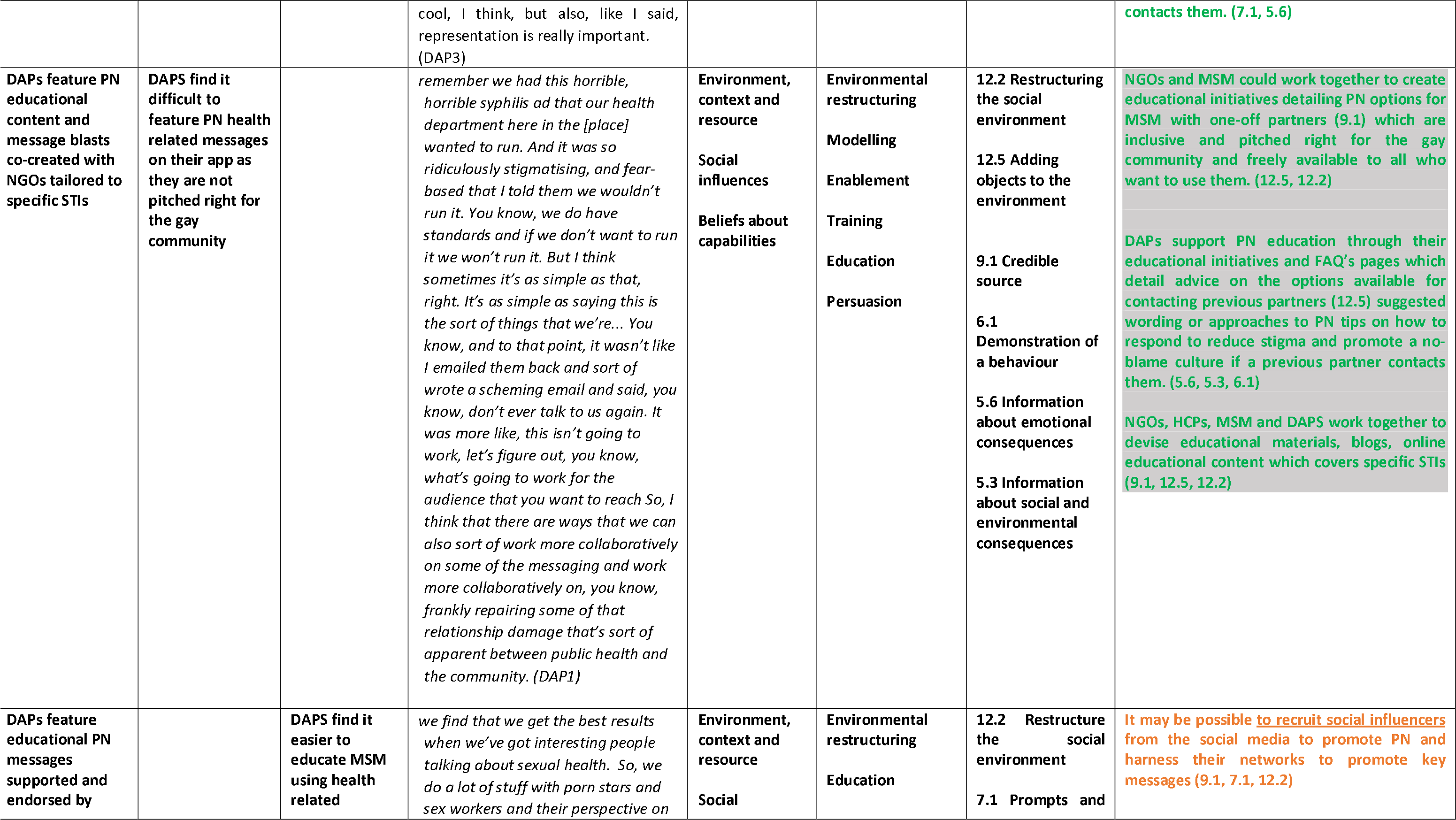
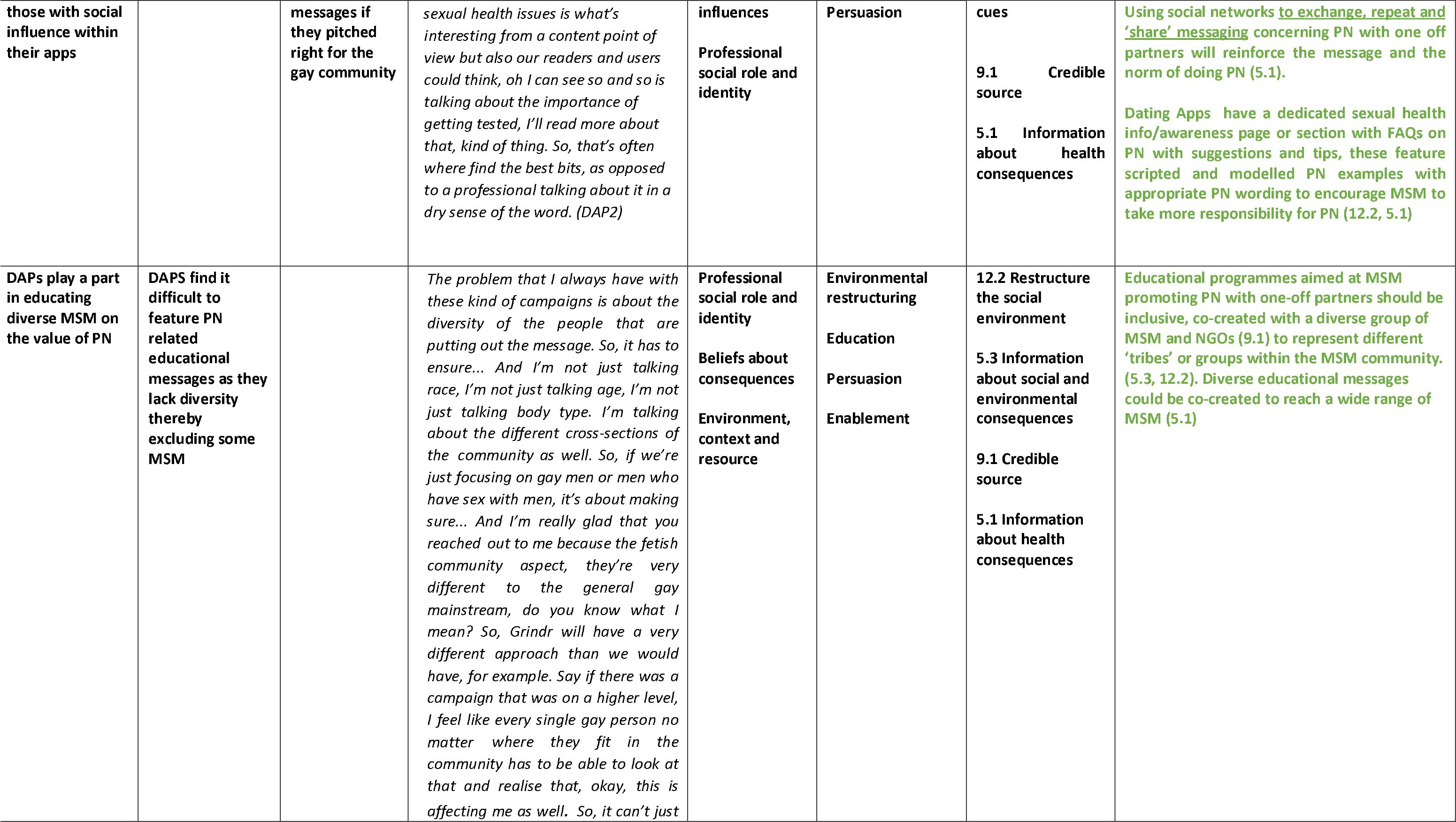
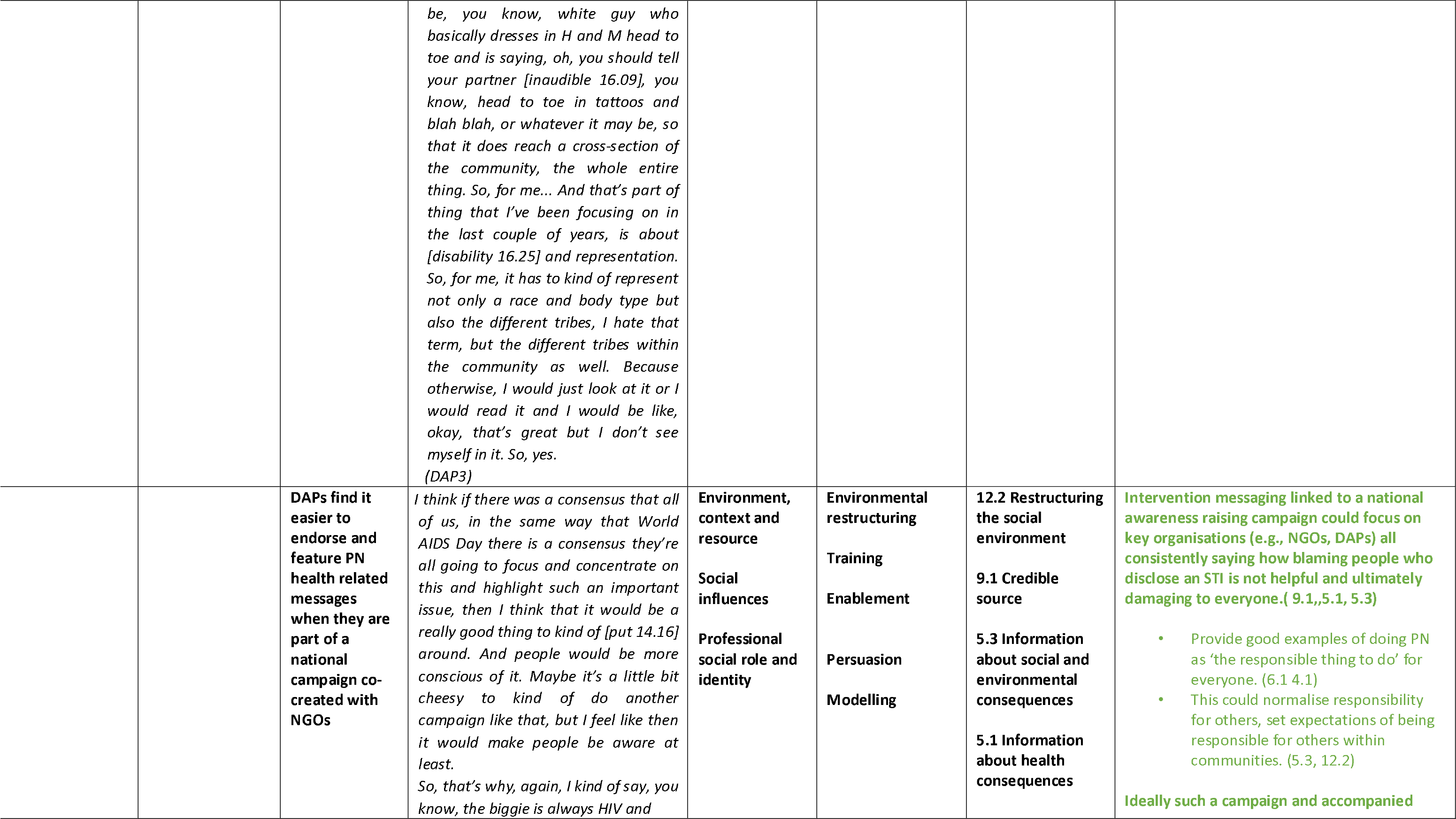
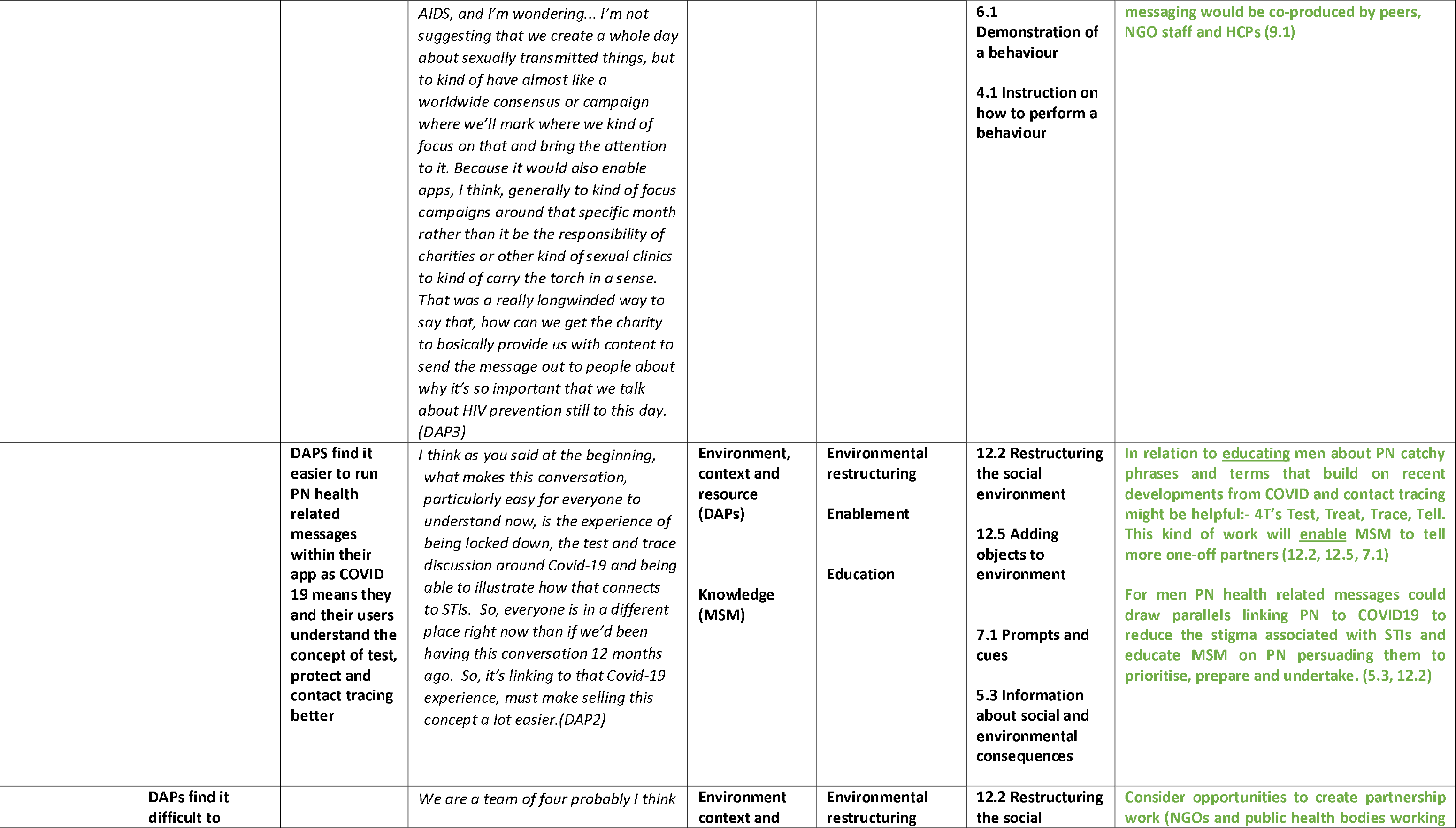
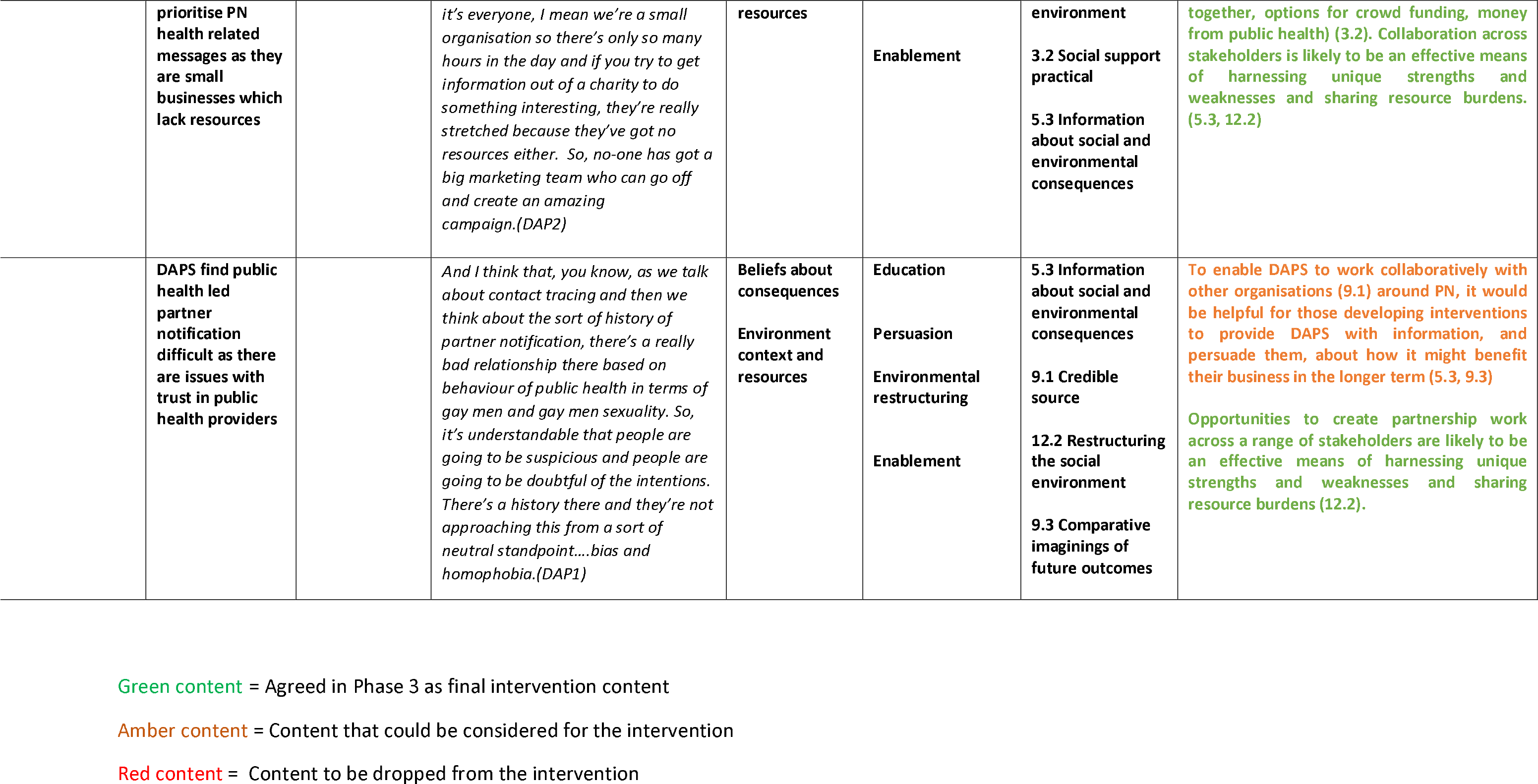
What did Dating app providers tell us about **implementing changes relating to promoting the endorsement of contact tracing within their infrastructure?**

❏ DAPs should host educational content on contact tracing on their platforms and website - developed in partnership with CBOs, GBMSM, HCPs and other key stakeholders - and ensure education is inclusive to GBMSM from all backgrounds. Educational messages can be supported, endorsed and amplified by those with social influence across other social media platforms. DAPs should consider building educational messages from recent developments in COVID-19 contact-tracing phrases and terms familiar to the public.
❏ DAPs should consider collaborations with social influencers to promote contact tracing educational content. DAPs should ensure content and messaging is linked to relevant national awareness raising campaigns, where possible. DAPs should consider opportunities for partnership and collaboration across stakeholders such as CBOs and public health bodies to harness strengths and share resource burden.
❏ Those aiming to collaborate with DAPs to promote contact tracing should provide DAPs with persuasive information on how education and advocacy will benefit their future work and positive experience of app users.

What did dating app providers tell us about **implementing changes relating to changing dating app functionality?** (Table 5)

**Table 5.**
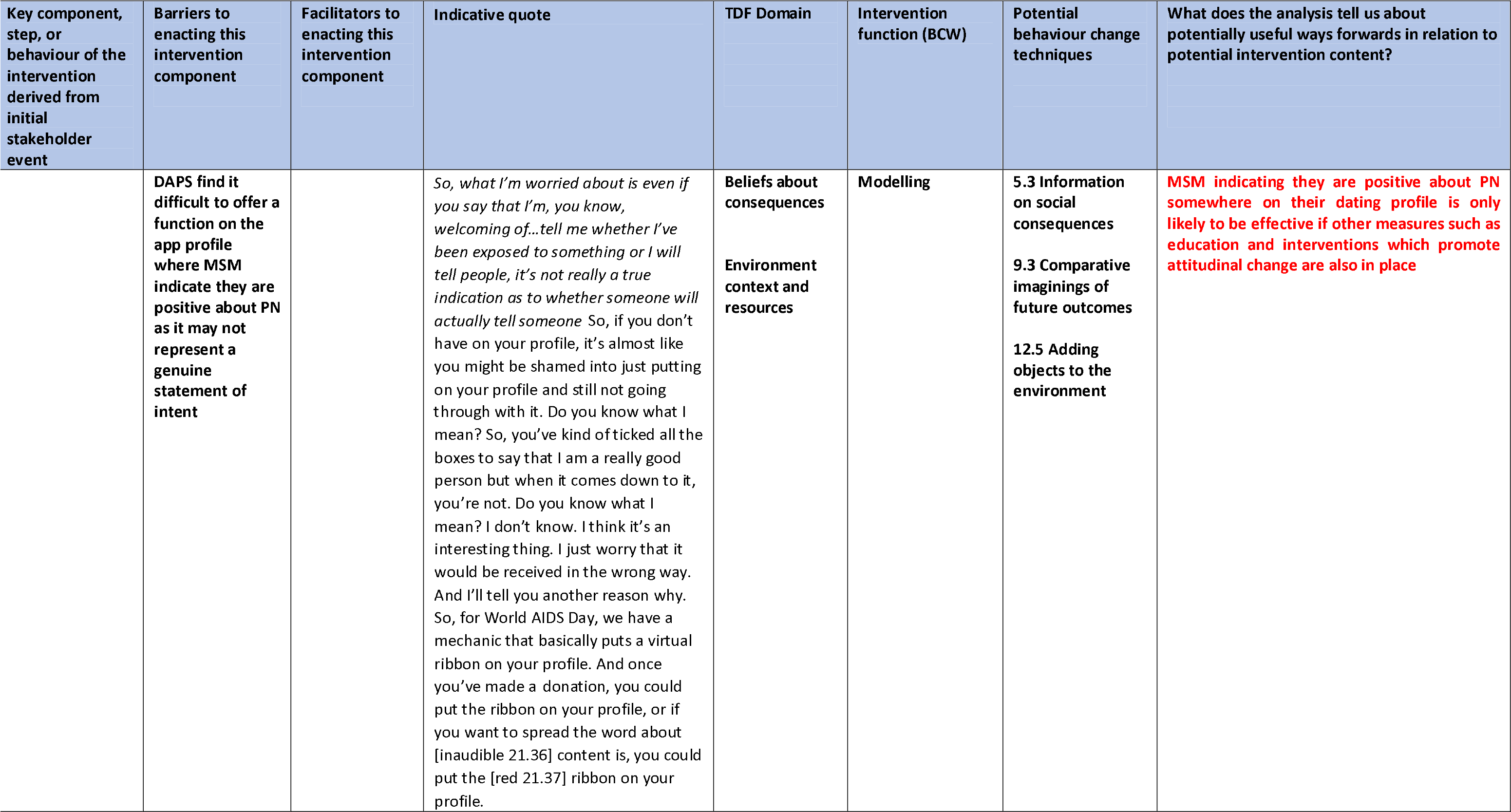
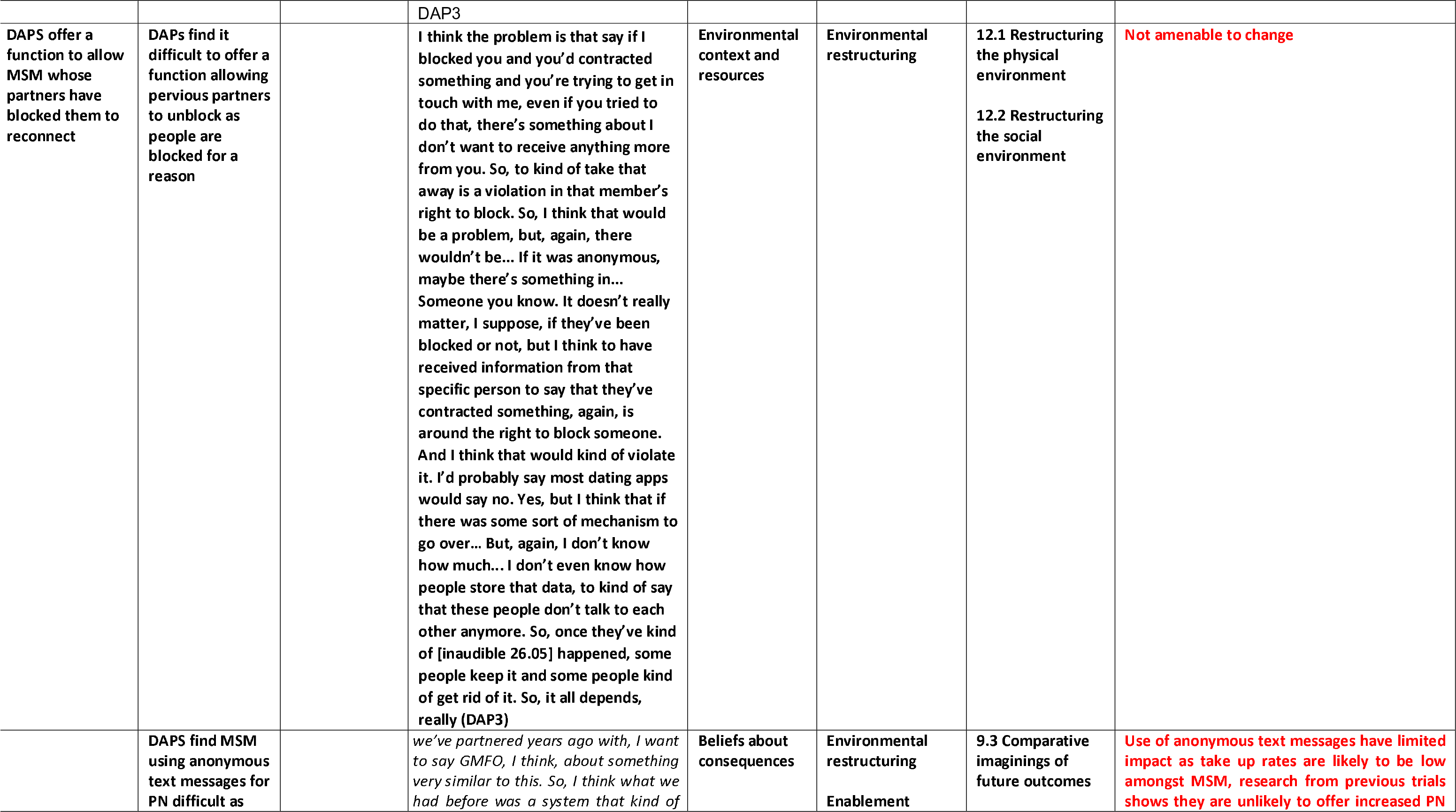
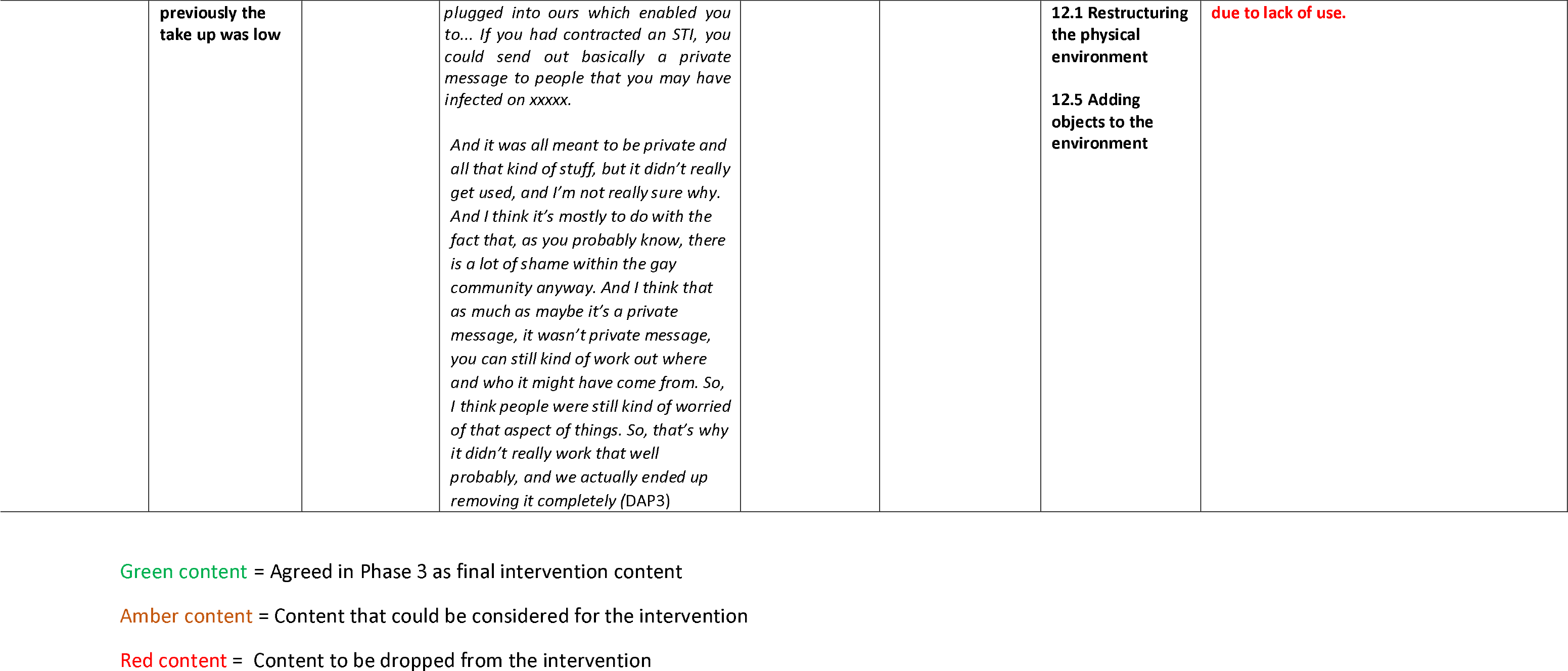
What did Dating app providers tell us about implementing changes relating to changing dating app functionality?

The intervention development process did not agree on recommendations for DAPs to implement changes to dating app functionality. DAPs were critical of developing in-app functions to enable users to indicate they are positive about contact tracing as this may not represent a genuine statement of intent and may infringe upon users rights. DAPs also indicated that such functionality would only prove effective if other measures such as education and interventions which promote attitudinal change amongst GBMSM are also in place. DAPs also suggested limited impact of functions which would enable anonymous contact tracing messages as uptake of this option is likely to be low amongst GBMSM; research from previous trials shows options for anonymous messaging are unlikely to increase rates of contact tracing due to lack of use. Finally, DAPs were also reluctant to develop functions which would enable users to ‘unblock’ former sex partners in order to send a message for contact tracing purposes, as this could be considered a violation of a user’s ‘right to block’.

What did dating app providers tell us about **implementing changes relating to signing up to a code of conduct to endorse contact tracing?** (Table 6)

**Table 6.** What did Dating app providers tell us about implementing changes relating to signing up to a code of conduct to endorse contact tracing ?

| Key component, step, or behaviour of the intervention derived from initial stakeholder event | Barriers to enacting this intervention component | Facilitators to enacting this intervention component | Indicative quote | TDF Domain | Intervention function (BCW) | Potential behaviour change techniques | Suggestions for intervention content |
| --- | --- | --- | --- | --- | --- | --- | --- |
| DAPs agree and sign up to a code of conduct supporting partner notification |  | **DAPs find it easier to sign up to a code of conduct if the right organisations work together** | A code of conduct around partner notification, you would need someone to bring it together, in terms of a charity working in the sector or someone who could get the right people at the table and then you could possibly...I would keep it really simple, I mean the challenge would be agreeing what are the standards that we're going to offer to users and what's the technology required to get there. Everyone would have to deal with their own technology because everyone runs their own platforms. So, it would be fairly broad and fairly general, a statement of intent, as opposed to anything else, I imagine. you could even do it where you can sign up to undertake to deliver opt-in partner notification by 2027 or whatever, that kind of thing. So, you<br>commit to being on a road map<br>towards enabling that<br>notification.(DAP2) | **Environment context and resources**<br>**Professional social role and identity**<br>**Reinforcement** | **Environmental restructuring,**<br>Persuasion<br>Enablement<br>Incentivisation | **12.2 Restructuring the social environment**<br>**3.2 Social support (practical)**<br>**12.5 Adding objects to the environment**<br>**1.8 Behavioural contract** | **NGOs MSM and public health organisations Lobby for code of conduct for dating app providers supporting PN. (3.2, 12.2)**<br>**Consider the development of an NGO and dating app led code of conduct (12.5). Key actors, i.e. NGOs, DAPs agree standards and co-produce a DAP code of conduct to support partner notification (12.5, 1.8)** |

Green content = Agreed in Phase 3 as final intervention content
Amber content = Content that could be considered for the intervention
Red content = Content to be dropped from the intervention

**Table 7.**
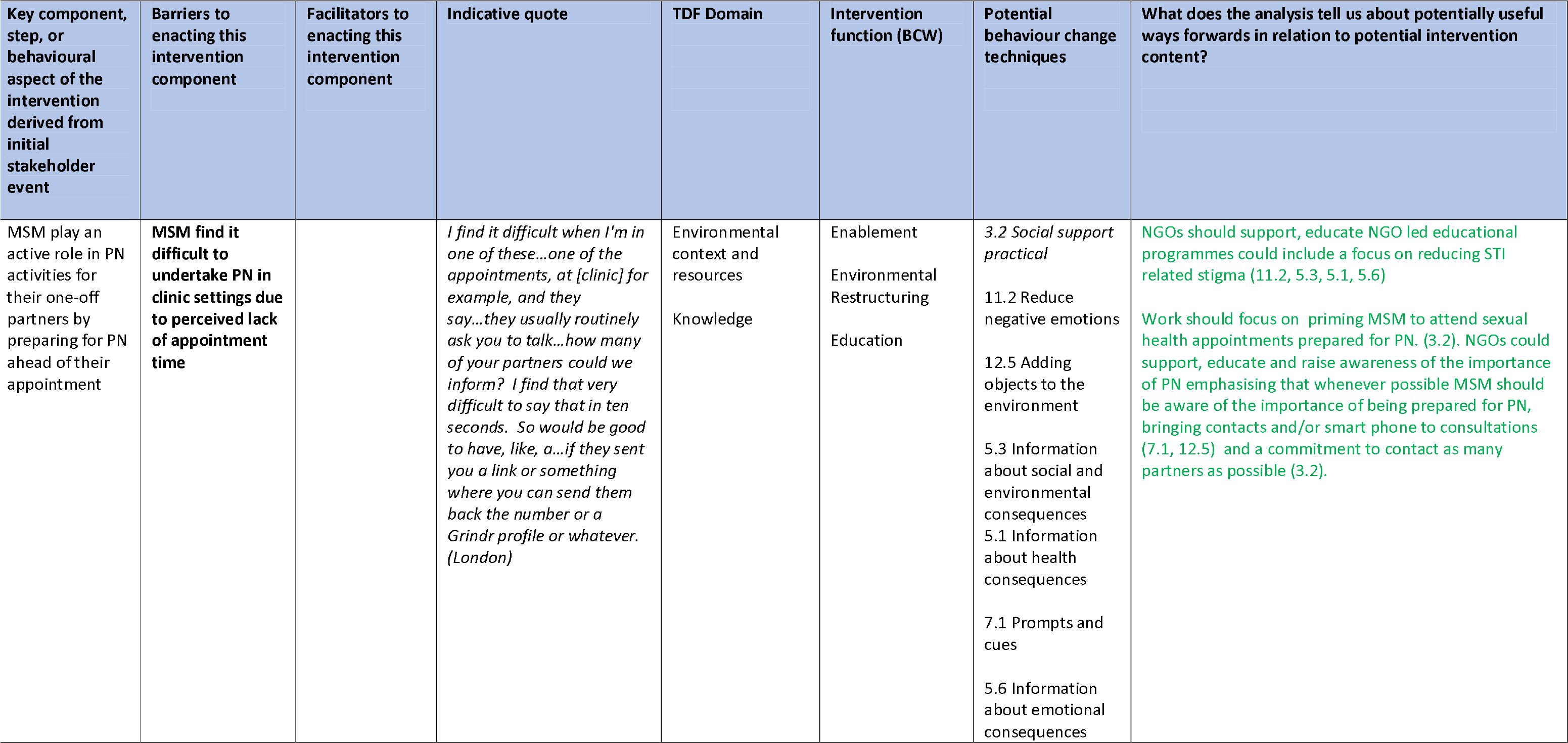
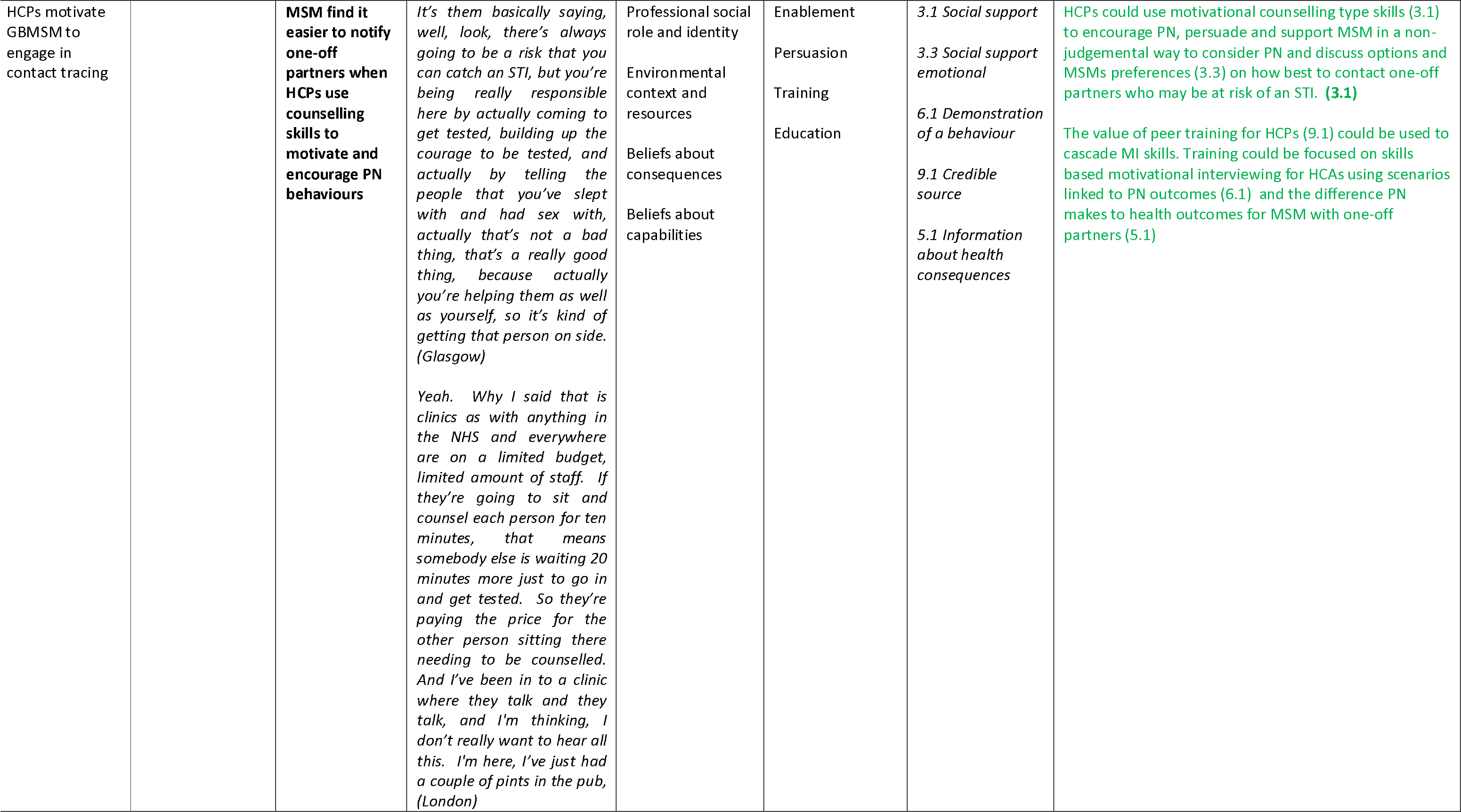
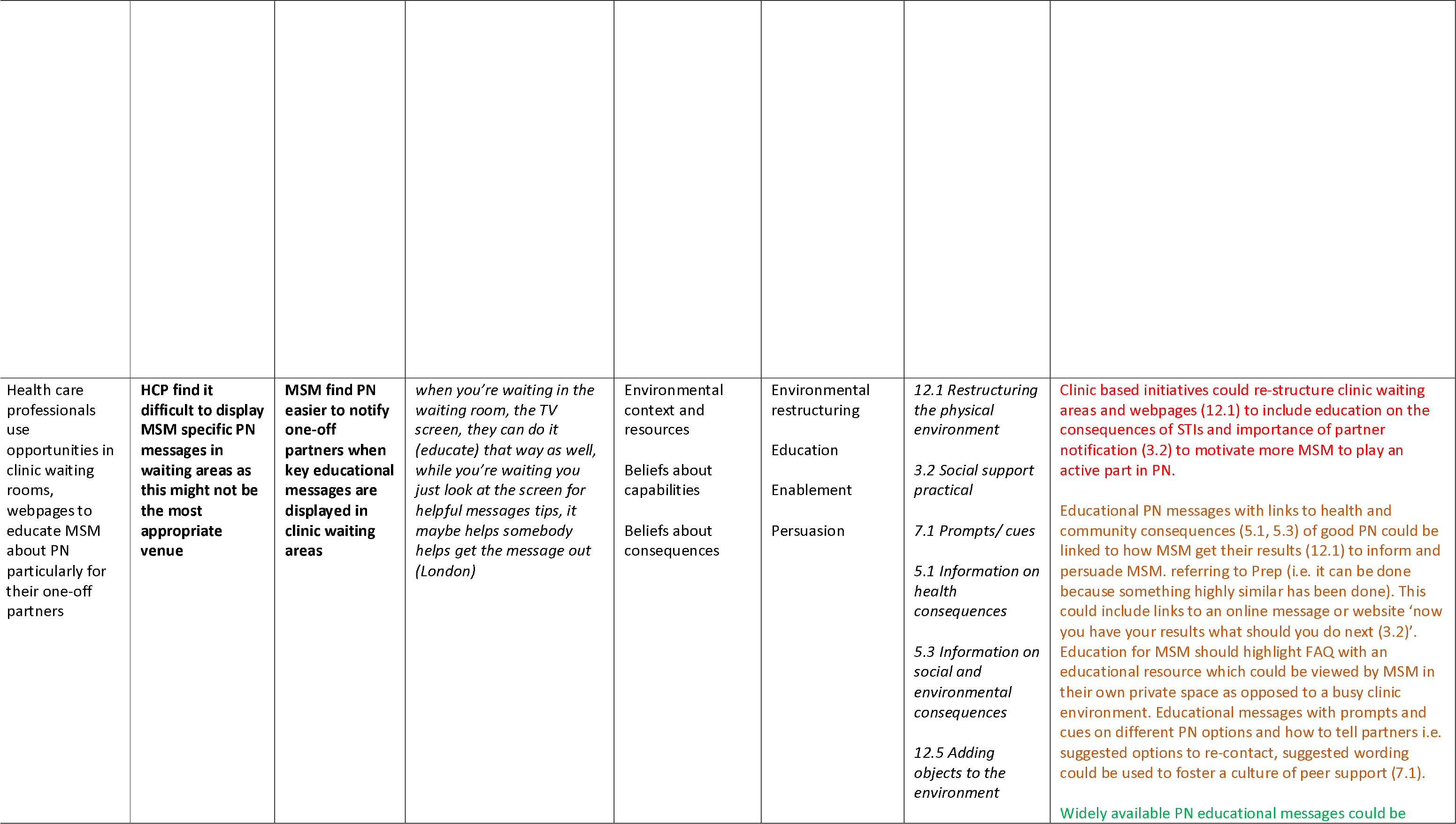
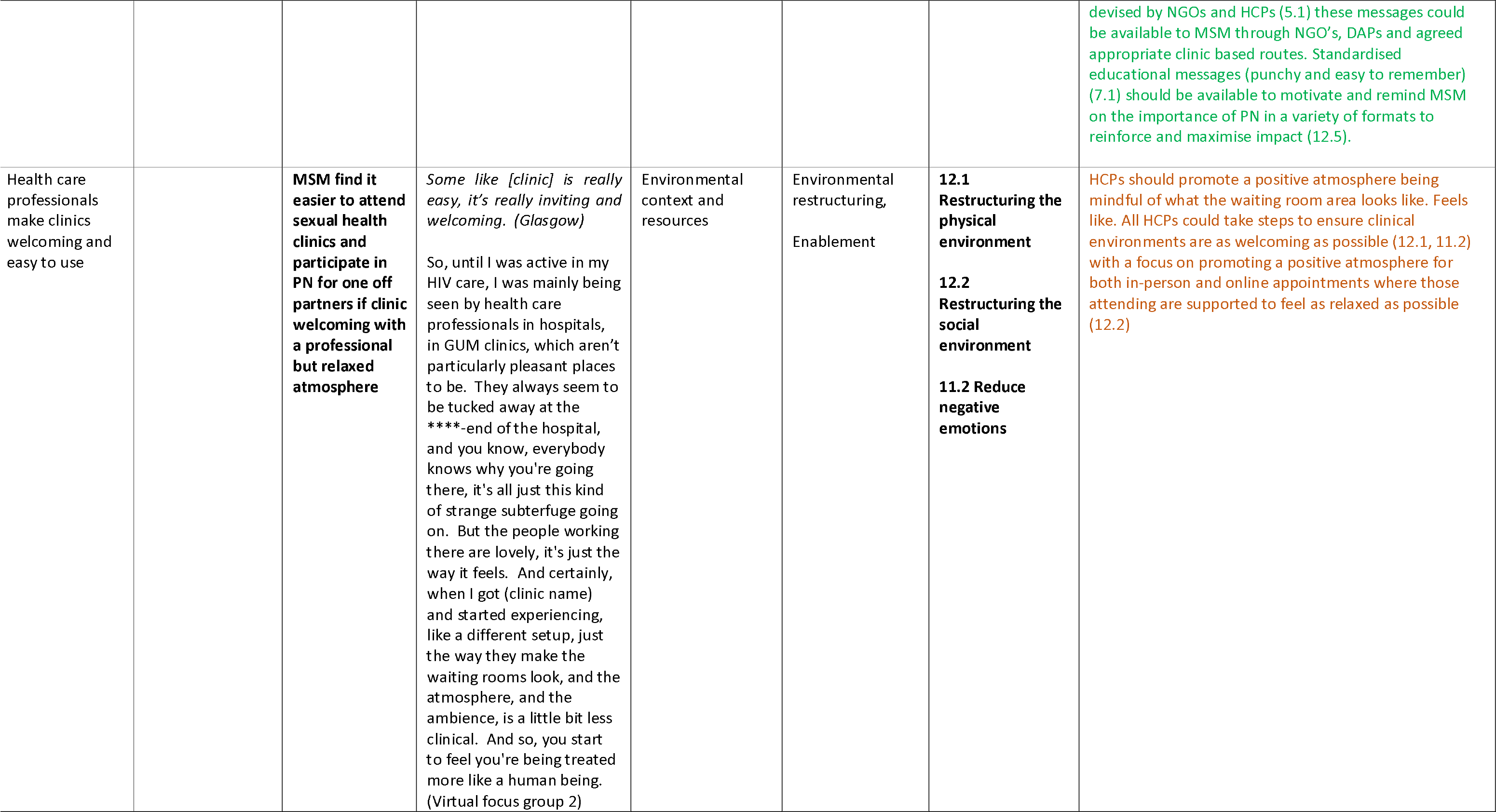
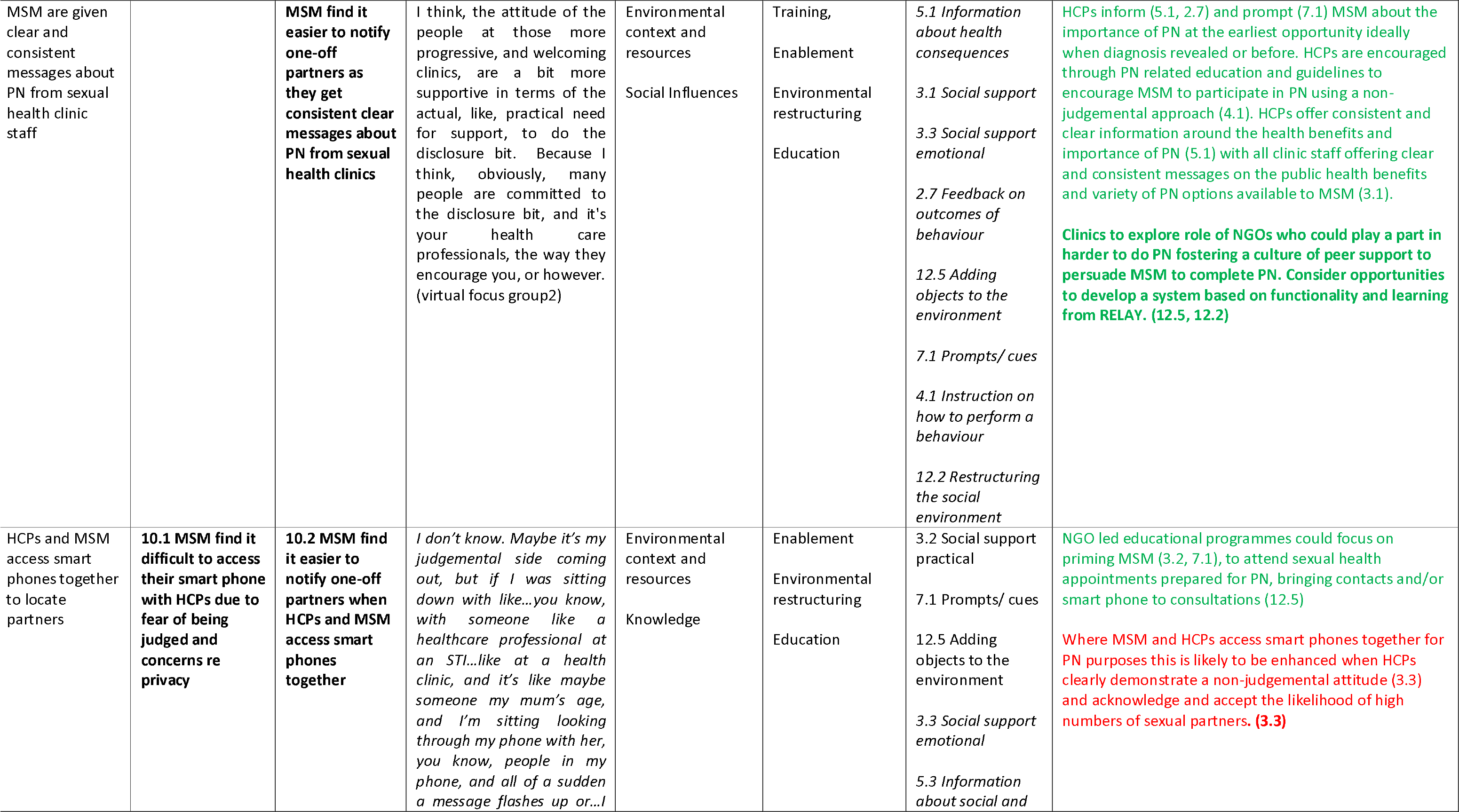
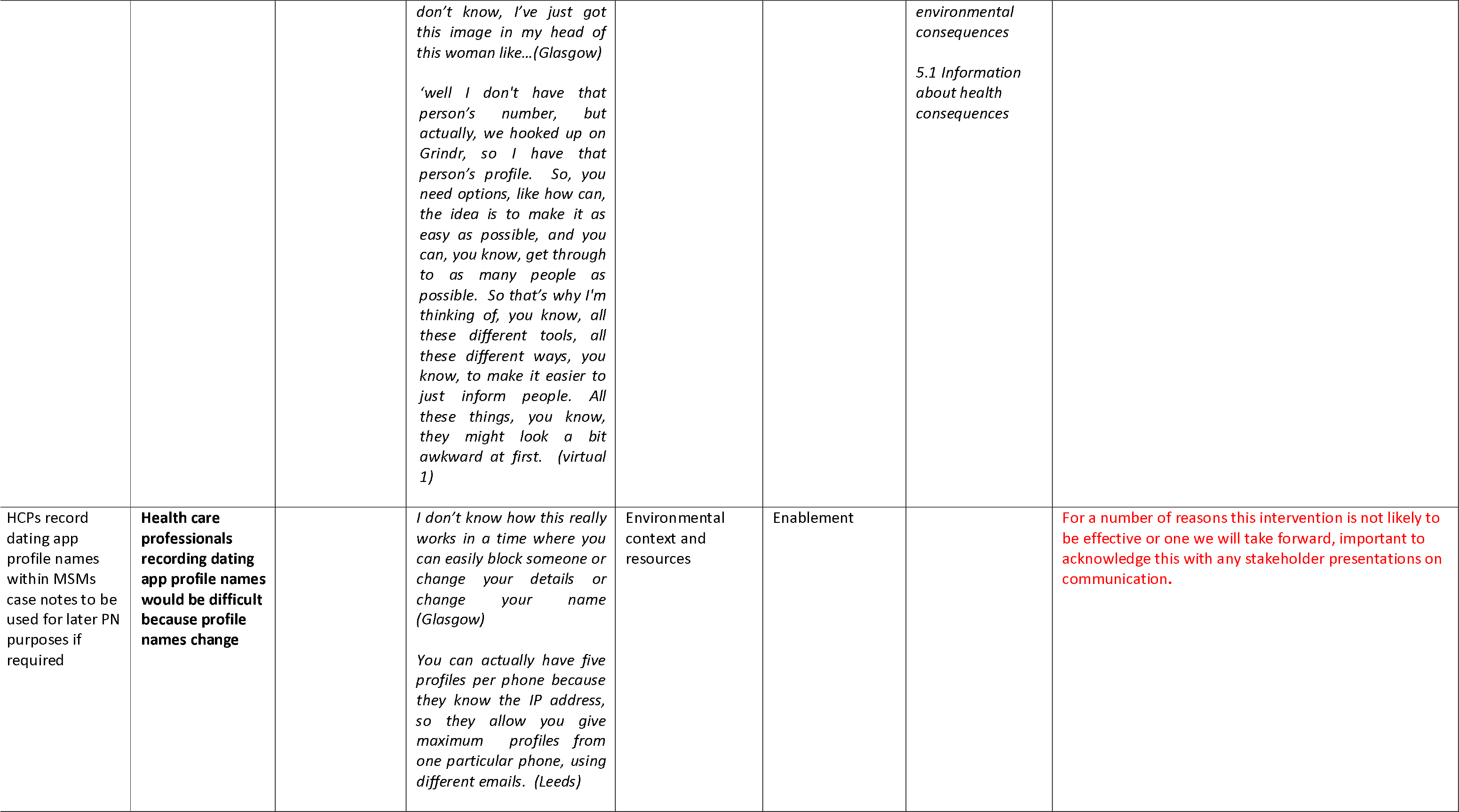
What have we learned about implementing improved interactions between HCPs and GBMSM to enhance contact tracing for one-off partners

The intervention development process did not agree on the recommendation for DAPs to sign up to a code of conduct supporting contact tracing, though the idea was not outrightly rejected. Stakeholders indicated that in order to be successful, a code of conduct would require lead from relevant organisations in development and encouraging commitment from DAPs. The following may be considered to enable this alternative approach.

❏ CBOs, GBMSM and public health organisations could lobby for a code of conduct for DAPs to endorse contact tracing. DAPs could work together with organisations to develop standards and co-produce a code of conduct for DAPs to endorse contact tracing.

What have we learned about implementing improved interactions between **HCPs and GBMSM to enhance contact tracing for one-off partners?** (Table 7)

❏ HCPs and sexual health clinics should form and utilise partnerships with CBOs and GBMSM to deliver culturally competent services. The delivery of contact tracing should be reframed to consider the roles of CBOs in supporting GBMSM and enabling them to play a more active role. Opportunities for partnership with CBOs should be explored to support over worked over-burdened clinic staff with contact tracing for GBMSM and their one-off partners. CBO educational initiatives could also promote the value and impact of individuals completing contact tracing themselves, as this helps to reduce stigma, create ownership and removes reliance on HCPs who have limited resources. GBMSM could co-produce materials with CBOs and communities to educate and enable other GBMSM to systematically raise issues of contact tracing with their HCPs
❏ Support GBMSM to prepare for contact tracing interactions with HCPs. HCPs should work to improve GBMSM engagement with contact tracing in-clinics through education, priming, and incentivisation. Educational messages for GBMSM should be devised by CBOs and HCPs and made widely available to GBMSM through CBO’s, DAPs and agreed appropriate clinic-based routes. HCPs should prime GBMSM at the earliest opportunity to enable and motivate them to participate actively in contact tracing, saving valuable clinic time and resources. HCPs should approach all contact tracing related activity in a non-stigmatising way, using guidance and training on how to perform this behaviour. Sexual health clinics should explore the potential role of CBOs in fostering a culture of peer support to persuade GBMSM to contact previous sexual partners in more difficult situations. Sexual health clinics should also consider opportunities to develop an online system for contact tracing, based on functionality and learning. Education for GBMSM should highlight FAQ and educational resources available for GBMSM to view in their own private space as opposed to a busy clinic environment. Educational messages could be linked to how GBMSM get their results to inform and persuade GBMSM to undertake contact tracing, as with similar sexual health interventions such as PrEP.
Education and training should be developed for HCPs on the value and importance of contact tracing, and to underscore promotion of culturally competent contact tracing as a key part of HCPs role. Promotion of contact tracing should be communicated as a key part of the role of HCPs and built into clinic standards, staff training and induction processes. Clear messages should be targeted to HCPs, developed in collaboration with CBOs, GBMSM and other relevant professional bodies. Clear and consistent messages on the value and importance of contact tracing should be communicated from credible sources such as reputable professional bodies (i.e. BASHH), commissioners, executive team members, senior clinicians, and consultants. Educational initiatives should be developed and delivered to HCPs, focusing on the consequences of poor contact tracing and the potential impact of the process. Educational initiatives should be planned and coordinated with a range of actors, and reinforce the positive public health consequences of contact tracing (i.e. CBOs, DAPs, sexual health organisations, and public health bodies). Educational initiatives should include understanding of GBMSM sexual cultures, to improve contact tracing for GBMSM with one-off partners. Skills-based training should be delivered to HCPs offering advice for consistent and effective contact tracing practice, and focusing on increasing HCPs knowledge and confidence to talk to GBMSM about contact tracing in the context of their sexual cultures.
❏ National networks, professional bodies and senior leaders should enable prioritisation of contact tracing through standardisation of contact tracing approaches, incentivisation, and training for HCPs. Clinic based consensus could be used to ensure a consistent approach towards contact tracing is adopted, with intervention approaches documented in clinic policies and procedures. Where possible provide prompts and cues these could be used with in-built algorithms on clinic computer system, clinic paperwork, posters around clinic areas, above desks with prompts and reminders about PN to motivate and remind HCPs of their PN role. Specific clinic initiatives and approaches to contact tracing should be standardised and clearly defined with prompts such as a flow chart and predefined questions to minimise demands on HCP time in light of resource constraints. HCPs should use a combination of professional judgement combined with guidance, hints and tips on good practice. Targets and goals related to contact tracing should be used as components of staff supervision and appraisal systems in clinics to incentivise contact tracing amongst HCPs.
❏ Commissioners and senior HCPs should consider in-clinic priority changes in relation to contact tracing and its value for public health. Links could be made to COVID-19 contact-tracing to stress the importance and value of increasing the percentage of partners contacted (i.e if 10% are reached, versus 50% or 80% of partners). Sexual health clinics should consider the value of all staff completing recognised training such as accredited CPD training with a focus on consistent terminology and phrases that can be used in contact tracing consultations tailored to GBMSM with one-off partners. More experienced staff could offer support and supervision through shadowing, live supervision as a method to boost confidence amongst other staff when asking about sexual cultures and documenting GBMSM partner types.

