## Supplementary File A for "Developing a co-produced, systems-informed, sexually transmitted infection contact tracing intervention for gay and bisexual men who have sex with men and their ‘one-off’ sexual partners"

*Distal drivers of contact tracing*

Across the small groups, there was clear consensus that more distal issues driving contact tracing were important, that the socioeconomic context of service provision and major elements of culture were vitally important to shaping contemporary contact tracing for one-off partners. Several groups highlighted contact tracing was shaped by the historic legacy of inadequate sex and relationship education which in turn was led by heterosexism and homophobia. In the future new kinds of wholistic compensatory sexual health education were needed to directly address contact tracing setting expectations, norms and values for life.

Most stakeholders acknowledged the enduring negative effects of a long history of pathologizing and blaming GBMSM, the problematic aspects of the business models of dating app providers, and the overstretched nature of contemporary sexual health services. All were seen as important in shaping contemporary contact tracing with one-off partners. Given the distributed nature of the drivers of contact tracing across the small groups, it was also felt that the burden of improving contact tracing should not fall on any one group of stakeholders alone. Instead, collective, and co-ordinated effort across the whole system would be helpful. Stakeholders talked of the potential value of saturating the system with clear signals always promoting and endorsing contact tracing, using peer interactions, websites, dating apps and a range of non-sexual health services (from mental health to emergency services) as well as contact tracing being a clear and consistent component within sexual health services themselves. There was particular enthusiasm for the idea of multiple stakeholders coming together to develop intervention components through the mass and social media to change GBMSM cultures and norms by actively promoting contact tracing, particularly ideas of integrating such messaging with dating app providers ‘message blasts’.

Most groups also discussed aspects of GBMSM cultures and communities which pose challenges for effective contact tracing. Against a backdrop of institutionalised homophobia and heterosexism, some aspects of GBMSMs cultures and communities were understood as problematic for contact tracing for one-off partners in particular. Other aspects were seen as key assets (i.e., powerful existing norms to engage in contact tracing amongst some GBMSM). It was felt that to improve contact tracing for one-off partners it was vital to first address its upstream and clearly social determinants; by using peer influence, by challenging sex- and STI-related stigma within GBMSM communities and by building community resilience. Small groups agreed on the importance of working with GBMSM to develop healthier ways of GBMSM interacting with each other, for example, collectively normalising responsibility alongside pleasure, with an emphasis on self-care as well as collective care.

A further part of the cultural drivers of contact tracing was the social world of dating apps; whilst partly reflecting the on-going and emerging sexual cultures of GBMSM the dating app industry also shaped them in ways which were not always conducive to contact tracing (e.g., ‘blocking’). These could be compensated for by DAPs adding new functionality (e.g., enabling app-users to consent to being unblocked for contact tracing) and clearly promoting and endorsing new norms for contact tracing.

*Proximal drivers of contact tracing*

Stakeholders also discussed how in addition to addressing the more ‘upstream’ drivers of effective contact tracing, a series of more ‘downstream’ drivers might also be important and were potentially amenable to change.

Across all the small groups, stakeholders drew attention to perceived lost opportunities and various new solutions to working within sexual health services to enhance contact tracing. GBMSM talked of how services were sometimes experienced as unwelcoming, and furthermore interactions with HCPs were perceived as judgemental and unhelpful. Contact tracing was inconsistently mentioned and sometimes not explained. Collectively stakeholders outlined a range of ways in which sexual health services could think about new ways of working to facilitate more contact tracing amongst GBMSM. Against a backdrop of resource dilemmas across sexual health services, stakeholders discussed the need for more opportunities to reflect on current and optimal practice – sharing what works with whom to continuously improve. Data driven approaches were mentioned as necessary for this work –for example, improving our understanding of current successes and failures through audit and monitoring, increasing effective collaboration with the third sector, bespoke training to ensure universal cultural competence for GBMSM and their sexual cultures, the development of conversational scripts to motivate GBMSM to engage in contact tracing with one-off partners, ensuring contact tracing was part of every conversation, educating all GBMSM on anticipating and preparing for a contact tracing interaction when STI diagnoses were likely, and finally, incorporating more routine technology, such as smart phones and dating apps, into contact tracing work within interactions in service. Other insights were raised by one or two stakeholders, these included ideas such as better outbreak management approaches. Several of the small groups highlighted how intervening at the level of sex parties, sex clubs or public sex environments might be useful.
